# A multi-modal vision knowledge graph of cardiovascular disease

**DOI:** 10.1101/2025.07.17.25331359

**Authors:** Khaled Rjoob, Kathryn A McGurk, Sean L Zheng, Lara Curran, Mahmoud Ibrahim, Lingyao Zeng, Vladislav Kim, Shamin Tahasildar, Soodeh Kalaie, Deva S Senevirathne, Parisa Gifani, Vladimir Losev, Jin Zheng, Wenjia Bai, Antonio de Marvao, James S Ware, Christian Bender, Declan P O’Regan

## Abstract

Understanding gene-disease associations is important for uncovering pathological mechanisms and identifying potential therapeutic targets. Knowledge graphs offer a powerful solution for representing and integrating data from multiple biomedical sources, but lack individual-level information on target organ structure and function. Here we developed CardioKG, a knowledge graph integrating over 200,000 computer vision-derived cardiovascular phenotypes from biomedical images with data extracted from 18 diverse biological databases modelling over a million relationships. A variational graph auto-encoder was used to generate node embeddings from the knowledge graph, which were used as input features to predict gene-disease associations, assess druggability and propose drug repurposing strategies. The model predicted new genetic associations and therapeutic strategies for leading causes of cardiovascular disease which were also associated with improved survival. Candidate therapies included methotrexate for heart failure and gliptins for atrial fibrillation. Imaging enhanced the ability to leverage biological data for pathway discovery. These capabilities represent an important step toward using biomedical imaging to enhance graph-structured models for identifying treatable disease mechanisms.

## Main

Comprehension of gene-disease associations is crucial for deciphering the molecular mechanisms underlying various diseases and identifying potential therapeutic targets^1^. Knowledge graphs (KGs) have been used to systematically model and interrogate the biology regulating complex systems and diseases at multiple scales of organisation^2^. A KG represents real-world facts and semantic relationships in a graph structure comprising nodes and edges. Entities within a KG represent genomics, transcriptomics, proteomics, molecular functions, intra- and inter-cellular pathways, phenotypes, therapeutics, and environmental exposures^3^. To construct a KG, information is aggregated from curated databases, non-standardised repositories, and evolving ontologies^4^. Machine learning approaches are applied to map entities and relationships to a low-dimensional vector space that represents its semantic structure more efficiently for downstream prediction tasks^5^.

While KGs provide a comprehensive framework for predicting novel gene-disease associations and prioritising candidates for further investigation^4,6^, they lack individual-level phenotypes encoding target organ structure and function. In this work we introduce CardioKG - a knowledge graph that integrates rich computer-vision derived phenotypes of cardiovascular structure and function from biomedical imaging with data sourced from diverse biological databases. This approach leverages human “endophenotypes”, which are closer to the pathophysiology of disease than other observable traits, to improve the prediction of gene-disease associations. We achieve this through developing an embedding algorithm that preserves the intrinsic properties of nodes and their directional relationship. We assess the performance of this vision-augmented KG, using 21 different imaging traits along with data from 18 biomedical databases for the detection of gene-disease associations and drug repurposing with functional enrichment analysis to assess the prioritised genes’ role in critical biological pathways. This work shows how biomedical imaging possesses semantically-meaningful information in multi-modal graph-structured models of human disease for precision medicine applications.

## Results

### Study overview

The UK Biobank study recruited around 500,000 participants aged 40 to 69 years between 2006 and 2010^7^. A sub-study recalled participants for cardiac magnetic resonance (CMR) imaging and computer vision analysis was subsequently used to measure 21 image-derived phenotypes capturing dynamic structural and functional systolic and diastolic traits of the ventricles, atria and aorta^8^. For building the KG we selected 4,280 participants who had imaging and a diagnosis of atrial fibrillation (AF), heart failure (HF), myocardial infarction (MI), hypertrophic cardiomyopathy (HCM), or dilated cardiomyopathy (DCM) (Supplementary Table 1), as well as a healthy group of 5,304 participants to capture a range of phenotypic diversity. In total over 200,000 image derived phenotypes were used in the model. Results are presented for the three most prevalent diseases. Baseline characteristics of the selected participants are summarised in Supplementary Table 2. A validation group of 489 participants without imaging was used for assessing drug repurposing outcomes.

We constructed CardioKG, a knowledge graph that integrates cardiovascular image-derived traits with external biomedical databases to model the complex biological relationships between genes, diseases and phenotypes. We then trained a variational graph autoencoder (VGAE)^9^ to generate node embeddings from the resulting KG using an approach we developed to preserve directional relationships, enabling the application of machine learning models to more accurately predict gene-disease associations. Functional enrichment^6^ analysis was used to identify related molecular mechanisms, biological processes and pathways. A druggability analysis^10^ of the predicted genes evaluated therapeutic potential. Finally, we conducted a drug repurposing^11^ analysis to identify existing medications that could be repositioned to target the diseases of interest. Figure 1 shows the overall design of the study and Figure 2 shows how the participants were selected. Figure 3 shows how computer-vision derived phenotypes were connected to other entities in the KG.

**Figure 1.**
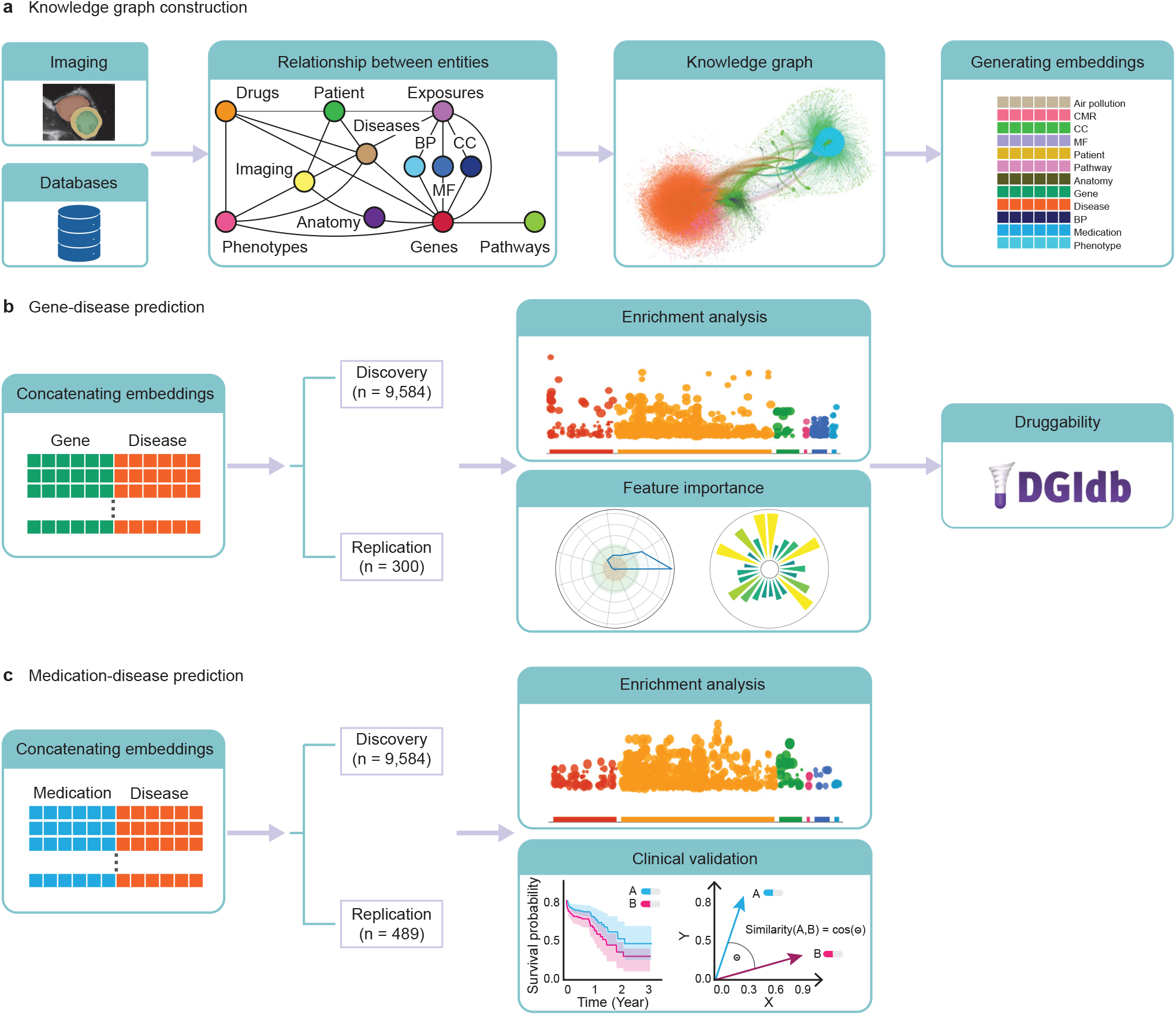
Overview of the knowledge graph construction and embedding process. **a**: Data were extracted from UK Biobank and 18 external databases to define entities (e.g., genes, diseases, medications, pathways, and imaging features) and their relationships. A schematic diagram illustrates the structure of the resulting multi-modal knowledge graph. A directed variational graph autoencoder (DVGAE) was then applied to learn low-dimensional embeddings for each node, preserving both topology and semantic relationships^4^. **b**: For predicting novel gene-disease associations we used a ground truth of known pairs from the DisGeNET^12^ database. Concatenated embeddings were used to train a machine learning classifier. Pathway enrichment analysis and ranking of feature importance was performed, and potential duggability assessed using DGIdb^14^. **c**: Using the same approach, known medication–disease pairs from the DrugBank^13^ database were used to predict new therapeutic associations. Validation was performed using enrichment analysis, outcome analysis, and graph-based methods. BP: biological process, CC: cellular component, MF: molecular function, CMR: cardiovascular magnetic resonance.

**Figure 2.**
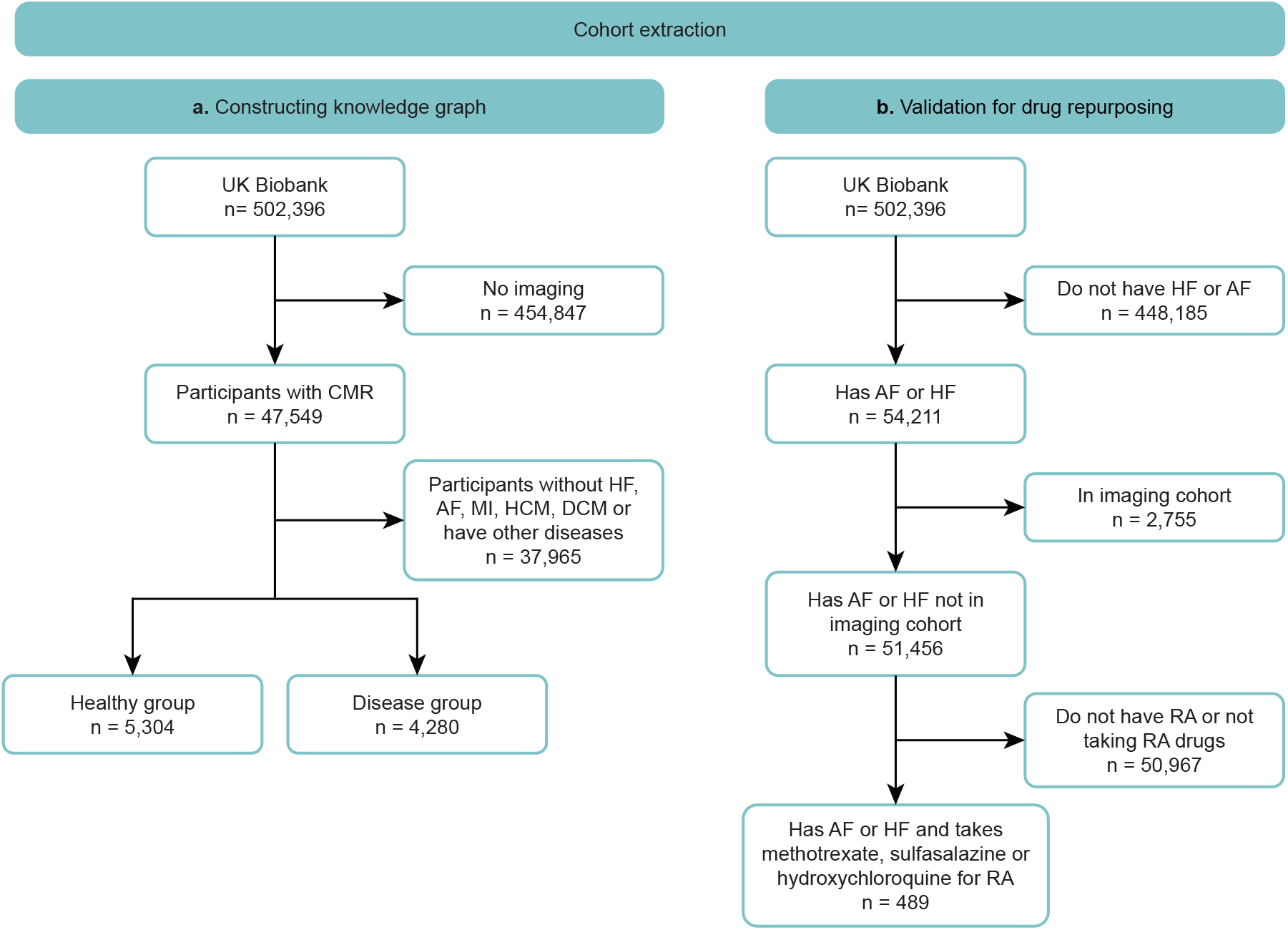
Cohort extraction from UK Biobank. **a:** A cohort (n = 9,548) to construct the knowledge graph included participants with atrial fibrillation (AF), myocardial infarction (MI), heart failure (HF), hypertrophic cardiomyopathy (HCM), dilated cardiomyopathy (DCM) and healthy individuals. **b:** A group (n = 489) to validate predicted medications using survival analysis in drug repurposing included those with AF or HF taking medication for rheumatoid arthritis. CMR: cardiac magnetic resonance, RA: rheumatoid arthritis.

**Figure 3.**
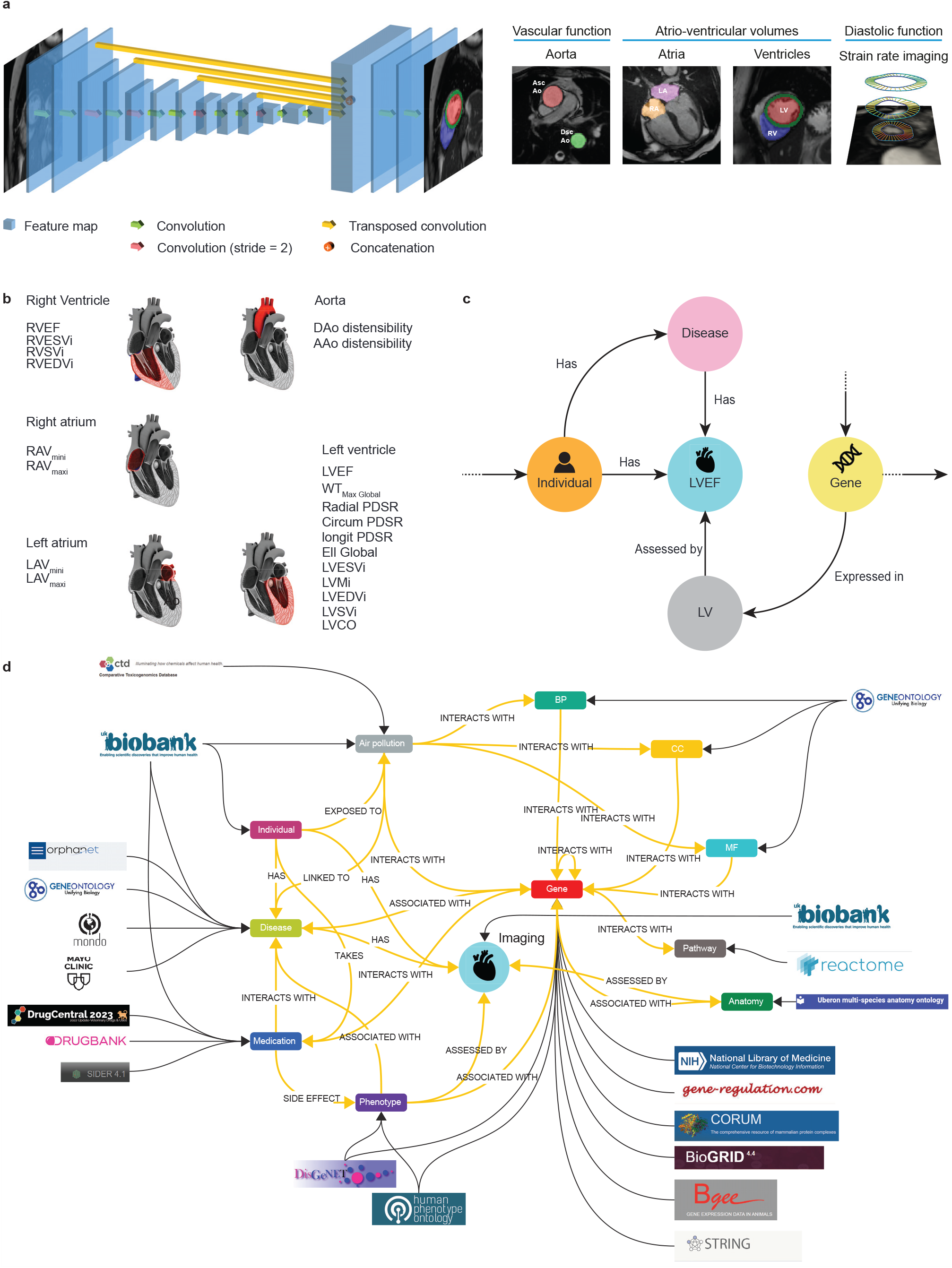
Integrating computer vision analysis to a knowledge graph (KG). **a**: A fully convolutional network processes cardiovascular magnetic resonance (CMR) images to output pixel-level segmentation maps. Motion analysis is also used to assess dynamic function of the heart and aorta. **b**: In total 21 imaging traits were derived assessing structural and functional traits in the atria, ventricles and aorta. The corresponding anatomical regions for each trait were represented as nodes in the KG, allowing linkage of imaging-derived features with related diseases and phenotypes. **c**: Imaging features are integrated in the KG as nodes. Each node color represents a specific entity in the KG linking imaging with diseases, anatomy, and gene expression. This panel displays a representative subgraph of the model, highlighting the integration of CMR-derived features within the KG. **d**: A schematic shows the relationships between the entities, the direction of the relationships and the source database for each entity. The imaging node is highlighted in the centre of the KG. RV: right ventricle, LV: left ventricle, LA: left atrium, RA: right atrium, Ao: aorta, AAo: ascending aorta, DAo: descending aorta, RAV: right atrium volume, RVEF: right ventricle ejection fraction, RVESV: right ventricle ejection systolic volume, RVEDV: right ventricle ejection diastolic volume, LVSV: left ventricle systolic volume, LVESV: left ventricle ejection systolic volume, LVEDV: left ventricle ejection diastolic volume, PDSR: peak diastolic strain rate, WT: wall thickness, RVSV: right ventricle systolic volume, LVM: left ventricle mass, LVCO: left ventricular cavity obliteration, LAV: left atrium volume, Ell: global longitudinal strain.

Ground truth labels for training a machine learning (ML) classifier to predict novel gene–disease and medication-disease pairings were obtained from DisGeNET^12^ and DrugBank^13^ databases. Associations not used in the construction of CardioKG were used for training. The embeddings were concatenated to preserve information from each model. Enrichment analysis was conducted for the top ten predicted genes, and we assessed the importance of related entities and imaging features. Druggability was evaluated using DGIdb^14^.

### Data extraction and knowledge graph construction

In addition to the imaging data we aggregated information from 18 biomedical databases (see Methods) to extract relevant knowledge on gene names, disease names, medication names, interactions between the medications and genes and other related biomedical terms, and converted the extracted data into a graph structure of nodes and relationships (Figure 3c). A total of 33,277 nodes and 1,195,437 relationships were extracted to construct CardioKG. The nodes included genes (n = 18,606), human participants (n = 9,584), medications (n = 2,106), and molecular pathways (n = 1,707). Phenotypic abnormalities were linked to human diseases (n = 1,036) and anatomical regions (n = 160) which included 5 structures segmented on imaging measuring 21 traits across each selected disease. Databases also included information on air pollutants, biological processes (BP), molecular functions (MF), and cellular components (CC).

### Embedding algorithm

KG embeddings offer a compact numerical representation of the graph’s structure and content enabling algorithms to learn and reason over the data. Each node (entity) and edge (relationship) is assigned a vector representation through minimizing a loss function. Here we developed an architecture based on a directed variational graph auto-encoder (DVGAE) which generates embeddings of the KG while preserving node and relationship properties as well as the directionality of relationships. The reconstructed graph accurately represented the original graph with high edge-wise accuracy of the embeddings (98.1%). Alternative methods such as Node2Vec^15^, TransE^16^, and ComplEx^17^ do not capture node and edge-specific features or directionality, which are attributes of our KG structure.

### Predicting gene-disease associations

Embeddings of genes associated or not associated with disease were concatenated with the embeddings of each disease of interest (Supplementary Tables 3 to 5). Ground truth labels for training used DisGeNET which is a database containing over 2 million gene-disease associations (GDAs), involving 29,000 genes and over 42,000 diseases with 4.3 million variant-disease associations (VDAs)^12^ aggregated from a dozen repositories that annotate clinically relevant variants (ClinVar) or genes (ClinGen, Genomics England PanelApp, among others). Three machine learning (ML) algorithms—random forest (RF), support vector machine (SVM) and artificial neural networks (ANNs), were trained on the concatenated embeddings to predict the association between the genes and disease using 5-fold cross-validation. SVM achieved the best performance based on accuracy (72.4% in HF, 75.0% in AF and 83.3% in MI), specificity (93.0% in HF, 97.0% in AF and 91.7% in MI), sensitivity (52.0% in HF, 51.6% in AF and 69.2% in MI) and area under the receiver operating characteristic curve (AUC-ROC,0.80 in HF, 0.78 in AF and 0.83 in MI) (Supplementary Table 6). Hence, SVM was used to predict novel associations for unlabelled genes without a known disease gene-disease association. Each predicted gene was assigned and ranked by a predicted probability of its association with disease. The top ten predicted genes (Supplementary Table 7) were selected as examples for functional enrichment analysis. A comparison was made for pathway enrichment using a KG without imaging traits. A hypergeometric test was used to determine whether a set of predicted genes is statistically overrepresented in a predefined reference gene set.

The top ten newly-predicted genes for HF included *GATA2, AGR1* and *EP300* which were significantly associated with 815 pathways (Supplementary Figure 1 and Supplementary Data File 1), among which 66 were identified as relevant pathways such as angiogenesis and the MAPK cascade (Supplementary Figure 2). These findings implicate genes in signaling cascades that regulate cellular regeneration and aging as potential genetic modifiers of HF. Variants in the MAPK pathway have also recently been implicated in GWAS data not used for KG training^18^. Without imaging traits only four relevant pathways were identified (Supplementary Figure 2 and Supplementary Table 7).

In AF, enrichment for prioritised genes including *SRC, GATA1* and *HSPA8* was identified in 658 pathways, of which 14 were relevant pathways associated with AF, including processes regulating cardiac conduction, response to hypoxia and regulation of immune system process (Supplementary Figure 3 and Supplementary Data File 2). These findings implicate genes that may regulate immune response and inflammation in AF, emerging as risk factors for arrhythmic disease^19^, and support a potential role for *SRC* which has been proposed as a promising target in other cardiovascular diseases^20^. Without imaging traits only one relevant pathway for AF was identified (Supplementary Table 7 and Supplementary Figure 3).

Lastly, in MI newly predicted genes included *PCNA, HTT* and *SNCA* which were significantly associated with 406 pathways, including 42 relevant pathways related to MI, such as apoptosis and cellular response to stress (Supplementary Figure 4 and Supplementary Data File 3). Enrichment analysis without CMR features revealed only four relevant pathways associated with MI (Supplementary Table 7 and Supplementary Figure 4).

### Druggablity of predicted genes

We assessed the therapeutic potential of the top ten predicted genes for each disease using the Drug-Gene Interaction Database (DGIdb) which curates information on drugs known to inhibit, activate, or otherwise modulate the activity of specific genes or their protein products. This evaluation focused on determining whether genes prioritised by CardioKG are actionable by existing drugs.

Among the top ten newly predicted genes associated with HF, five were identified as druggable (*AR, APP, GATA2, EGR1*, and *EP300*). These genes have been recognised as potential therapeutic targets and can be modulated by a total of 48 medications. Drugs including the monoclonal antibodies ponezumab and bapinezumab have been identified as potential candidates for targeting *APP* for instance (Supplementary Figure 5). For newly-associated AF genes, seven were druggable (*SRC, CASP8, DAPK1, H2AX, HSPA8, EP300, HNF4A*). These genes can be targeted by 37 medications including several anti-diabetic “gliptins” (dipeptidyl peptidase-4 inhibitors) which have observational evidence of a potential anti-arrhythmic role in treated diabetic patients (Supplementary Figure 6)^21^. Finally for MI, of the predicted genes two were identified as druggable (*SNCA* and *H2AX*). These genes can be targeted by 4 medications, including apoptosis inducers eltanexor and selinexor (Supplementary Figure 7).

Without CMR features in the KG only two genes were identified as potentially druggable for each of HF (Supplementary Figure 8), AF (Supplementary Figure 9) and MI (Supplementary Figure 7).

### Importance of image-derived phenotypes

The significance of encoded imaging traits was assessed by PageRank^22^, which quantifies node centrality based on the structure of incoming connections, assigning higher scores to nodes connected to other highly ranked nodes (Supplementary Figure 10). The CMR entity had the highest score (51.09 - 51.60) reflecting imaging’s central position within the graph and connection density to other node types (Figure 4) with left ventricular ejection fraction the highest ranked feature.

**Figure 4.**
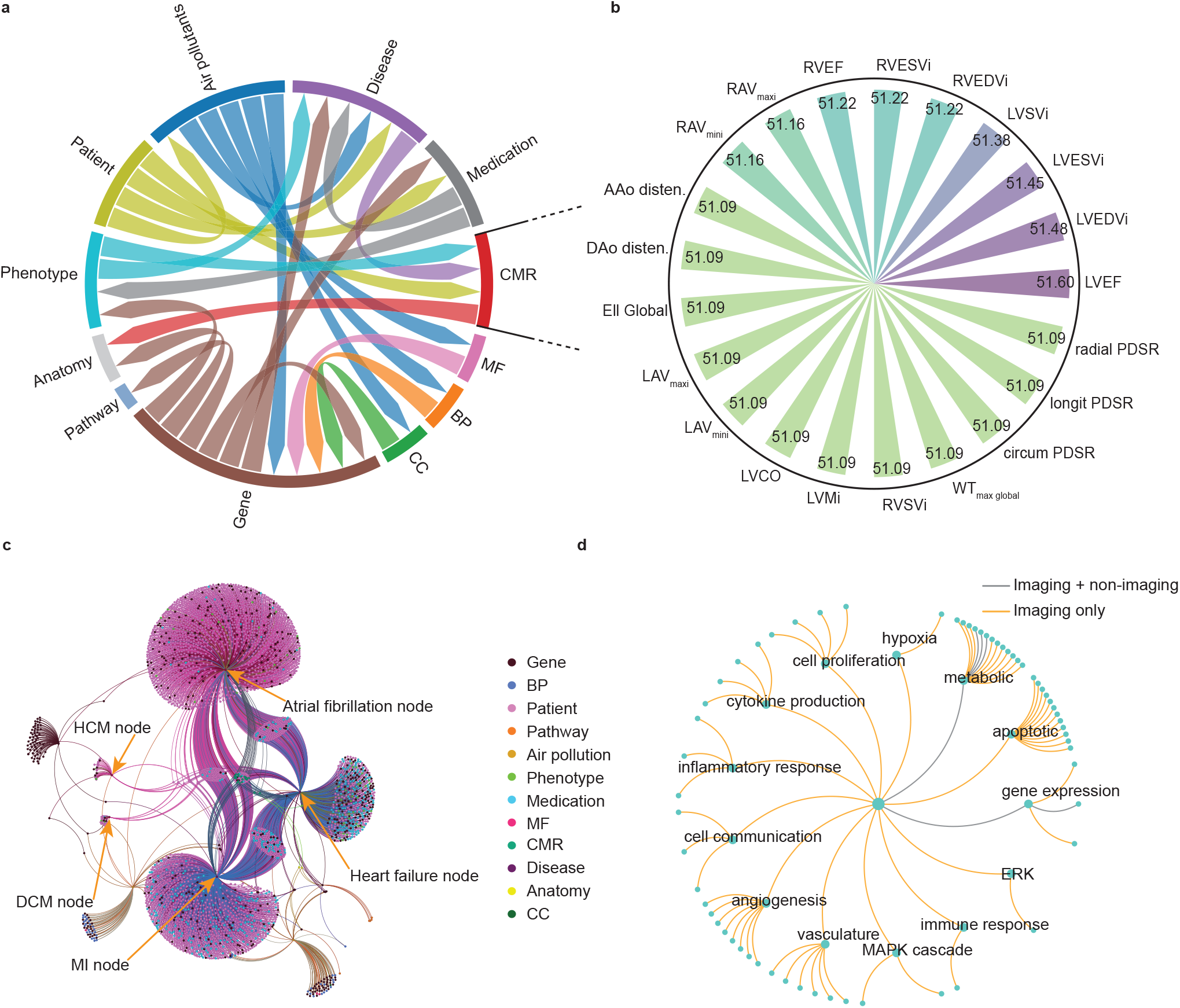
Contribution of cardiovascular imaging features to the structure and inference of the knowledge graph. This figure illustrates how cardiovascular magnetic resonance (CMR) features integrate into the knowledge graph and influence its structure and downstream analyses. The aim is to assess the relative importance of CMR-derived imaging parameters to evaluate whether including imaging improves pathway discovery. **a:** A circos plot shows the extracted entities and the directional relationships between them, serving as a compact visual summary of the underlying knowledge graph structure. **b:** PageRank scores for each imaging parameter (n=21) within the knowledge graph indicating their relative influence and connectivity to other influential nodes. Left ventricular ejection fraction ranked highest, followed by right ventricular and atrial parameters. Other values, including aortic function and strain shared similar scores. **c:** Network interactions among genes, diseases, pathway, medications, and CMR features. **d:** Enriched pathways identified with and without the use of imaging. Pathways highlighted in orange are specific to analyses using imaging data only, whereas those in gray represent pathways enrichment whether imaging data was used or not. HCM:hypertrophic cardiomyopathy, DCM: dilated cardiomyopathy, MI: myocardial infarction, MF: molecular function, CC: cellular component, BP: biological process, AAO: ascending aorta, DAO: descending aorta, RAV: right atrium volume, RVEF: right ventricle ejection fraction, RVESV: right ventricle ejection systolic volume, RVEDV: right ventricle ejection diastolic volume, LVSV: left ventricle systolic volume, LVESV: left ventricle ejection systolic volume, LVEDV: left ventricle ejection diastolic volume, PDSR: peak diastolic strain rate, WT: wall thickness, RVSV: right ventricle systolic volume, LVM: left ventricle mass, LVCO: left ventricular cavity obliteration, LAV: left atrium volume, Ell: global longitudinal strain.

To evaluate their structural role, we conducted an ablation experiment in which CMR nodes were removed from the graph. This resulted in decreased performance of the SVM model (Supplementary Table 6). The CMR nodes function as intermediaries linking anatomical features to genetic and disease-level data with their removal affecting graph connectivity which impairs the model’s capacity to capture indirect associations. This leads to predictions that are less aligned with disease-relevant pathways.

The number of relevant pathways identified for diseases of interest is also greater when CMR features are included (χ^2^,*P* = 0.001). Together these findings indicate that the inclusion of imaging nodes enhances the KG’s structural and functional complexity, enabling the model to better leverage interconnected biological data to uncover genes linked to a broader range of critical pathways.

### Drug repurposing

Embeddings of the diseases of interest and medications either indicated or contraindicated for them were concatenated and used to train a model to predict novel disease-medication associations. The predicted medications were further evaluated through enrichment analyses of their target genes, survival analysis and graph-based validation.

The KG-based machine learning model identified potential associations between HF and a range of existing medications. Among these, the top ten candidates included methotrexate, topiramate, and ranolazine prioritised based on their predicted association scores (Supplementary Table 8). Pathway enrichment analysis of the predicted medications was performed using known target genes. The target of methotrexate, *DHFR*, was linked to regulation of oxidative stress response which is a key factor in the pathophysiology of heart failure and cardiac remodeling^23^. Additionally, the targets of topiramate, including *SCN5A, SCN10A, CACNA1C*, and *CACNA1D*, showed significant associations with essential HF pathways, such as myocyte contraction and action potential regulation. The full enrichment analysis results are in Figure 5.

**Figure 5.**
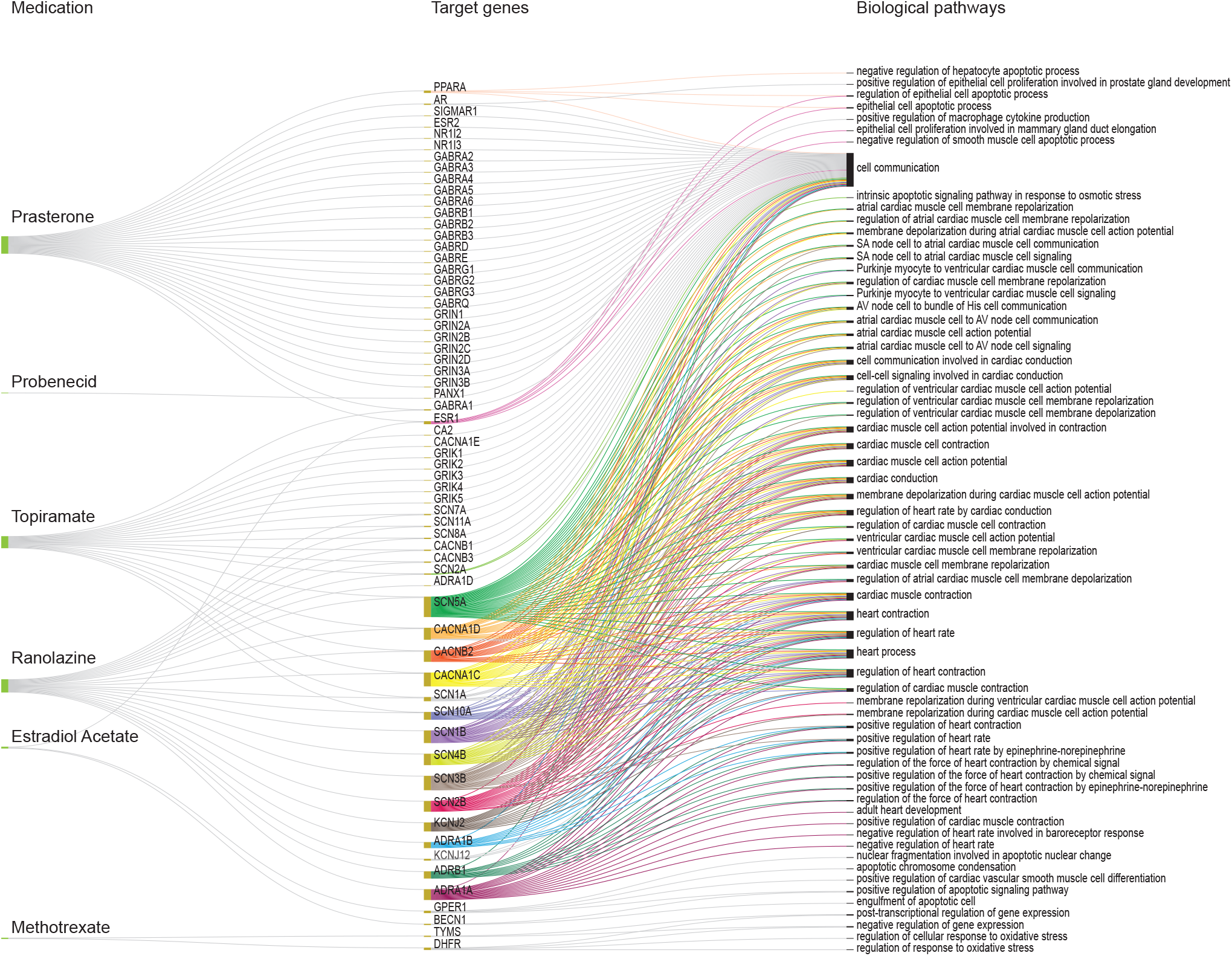
Enrichment analysis of target genes for predicted medications treating heart failure. Enrichment analysis was conducted to identify critical pathways for heart failure (HF) that are significantly associated with the target genes. The figure illustrates the predicted medications for HF, their associated target genes, and the enriched pathways linked to those genes. This figure excludes associated non-pharmaceutical substances (glutamic acid, caffeine, and cannabidiol). The same enrichment analysis was carried out for atrial fibrillation and myocardial infarction (Supplementary Figures 11 and 12).

A subset of UK Biobank participants, not included in model development, was used to determine whether any of the predicted medications were associated with improved survival outcomes in HF. Methotrexate was the only candidate included in the survival analysis due to the limited number of individuals exposed to most of the predicted medications. Here participants diagnosed with HF and rheumatoid arthritis (RA) were considered, since RA is a common indication for the use of methotrexate. Three subgroups were identified based on treatment with commonly prescribed RA medications:(1) patients with RA taking methotrexate (n = 147), (2) patients with RA taking sulfasalazine (n = 40), and (3) patients with RA taking hydroxychloroquine (n = 47) (Supplementary Table 9). All three medications are disease-modifying antirheumatic drugs (DMARDs) routinely used for the management of RA. Heart failure patients also being treated with methotrexate for RA had higher survival probability compared to those treated with hydroxychloroquine (hazard ratio (HR) = 1.76 [1.11, 2.79], *P* < 0.02) and sulfasalazine (HR = 1.63 [0.98, 2.72], *P* = 0.05) (Supplementary Figure 13 and Supplementary Table 10). The medications currently approved for HF and the KG-predicted medications had high cosine similarity scores (Supplementary Figure 14), which indicates similar vector representations, suggesting they share similar relational or structural contexts within the graph reinforcing their potential therapeutic relevance (Supplementary Figure 14).

A similar analysis was conducted for AF, where the top ten candidate medications, including methotrexate, zonisamide, acamprosate, and probenecid, were prioritised (Supplementary Table 11). Enrichment analysis revealed that the target of methotrexate, *DHFR*, is also involved in cellular pathways that promote AF^24,25^. Additionally, the targets of zonisamide, including *SCN5A, SCN4B*, and *SCN1B*, are associated with relevant pathways in AF, such as the regulation of cardiac muscle cell membrane repolarization and depolarization (Supplementary Figure 11). A similar survival analysis was performed in patients with both RA and AF being treated with methotrexate and alternative drugs (Supplementary Table 12) but the prevalence was low compared to HF and no significant differences in survival probability were observed (Supplementary Figure 15 and Supplementary Table 13). Network-based validation revealed that the predicted medications shared high cosine similarity with those already indicated for AF, supporting their therapeutic relevance and biological plausibility (Supplementary Figure 16).

For MI the ten highest-ranking drug candidates, selected based on their prediction scores, were prioritised (Supplementary Table 14) for pathway enrichment analysis in the same way. This included vorinostat which targets HDAC6 in the regulation of cellular responses to oxidative stress and the regulation of reactive oxygen species (ROS) metabolism (Supplementary Figures 12 and 17). Survival analysis was not performed as these drugs are not commonly prescribed. A graph-based validation also indicated high cosine similarity between the predicted medications and those already approved for MI (Supplementary Figure 18).

## Discussion

A KG is a semantic network representing the relationship between diverse real-world biomedical entities to enable systematic study of human disease. CardioKG provides a holistic view of cardiovascular disease by integrating data across 19 diverse biological databases representing relationships between genomics, molecular pathways, and exposures. Here we show how precision phenotypes, abstracted from computer vision analysis of the heart and circulation, leverage such rich interconnected biological data to uncover novel candidate genes and therapeutics linked to three major causes of morbidity and mortality worldwide^26^. We also demonstrate the potential of predicted drug repurposing to improve patient outcomes in heart failure. Together this shows how the performance of disease-specific semantic models can be substantially improved using cardiovascular imaging.

Genome-wide association studies find associations between common variants and disease,^27^ but have limitations in identifying causal mechanisms, rely on single traits and do not leverage prior knowledge. While recent efforts using machine learning, such AI-based summarisation, aim to improve GWAS interpretation^28^, most associated variants still have individually small effects on traits or diseases and rarely indicate actionable targets. In contrast knowledge graphs have emerged as a powerful framework for integrating data across multiple domains to predict novel gene-disease associations^29–31^. An important bottleneck has been the limited availability of individual-level phenotypes that can be linked to other semantic information in the network. Through leveraging advances in image segmentation and motion tracking in large biobank populations with MRI we show how a KG network can be enriched by quantitative organ phenotypes. In CardioKG a node for each image-derived phenotype is semantically linked to nodes encoding anatomical sites, genes and diseases. We show how this offers greater pathway enrichment for discovered genes, higher yield of potentially druggable targets, and biologically plausible predictions for drug repurposing. We also introduce a framework that generates KG embeddings while preserving directionality in node and relationship properties achieving high reconstruction fidelity.

Taking HF as an example, CardioKG prioritised genes linked to regulatory pathways that include apoptosis, angiogeneisis, inflammation and tissue hypoxia. One prioritized gene is *APP* which is associated with amyloidogenic pathways in Alzheimer’s disease, but this multi-organ condition is also associated with systemic inflammation and oxidative stress affecting peripheral organs including the heart - likely associated with *A*β amyloid deposition^32^. The KG predicted several pharmaceutical compounds interacting with *APP* as agonists, inhibitors, or modulators. Among these, the KG also predicted humanised monoclonal antibodies as potential therapeutics targeting *APP*^33,34^. The KG proposed methotrexate, topiramate, and ranolazine for drug repurposing in HF with enriched associations for myocardial contraction and action potential regulation. Methotrexate also showed a potential survival benefit in patients with both HF and rheumatoid arthritis compared to other treatments. A trial of methotrexate to evaluate its cardiovascular benefits in patients with RA is underway^35^ and it has a favorable repurposing safety profile and cost-effectiveness^36^. Its benefits are independent of reductions in RA disease activity suggesting the KG has identified alternative methotrexate-related mechanisms in modulating cardiovascular risk^37^. The KG also prioritised anti-epiletic therapies that target *SCN5A*-regulated ion channels in the conduction system as treatments for AF, and vorinostat as a repurposed treatment for MI which targets *HDAC6* regulation of mitochondrial biogenesis^38^.

Limitations of this work are that the UK Biobank population is primarily of European descent highlighting the need for enriching the KG image-phenotypes with more diverse populations. Technologies for acquiring knowledge from multiple sources and integrating them into a KG structure are still developing, and there are technical challenges for graph completion, knowledge fusion, and efficient reasoning that are areas of active development^39^. There are no established standards yet for graph-based biomedical data models but these may emerge as their use becomes more widespread. The use of large language models may complement the ability to acquire knowledge from unstructured text but on their own may give inaccurate or inconsistent results in biomedical knowledge discovery^40^.

Our findings point to the potential for vision-based KGs, that capture phenotypes closely coupled to underlying individual-level pathophysiology, to accelerate discovery of new therapeutics in cardiovascular science as well as medicine more broadly. Biomedical imaging encodes additive information in semantic networks and the versatile architecture of CardioKG is generalisable across multiple disease use-cases where imaging is available. Future opportunities may lie in personalising diagnostic strategies through leveraging larger and more diverse population datasets.

## Methods

The UK Biobank cohort study recruited over 500,000 participants aged 40 to 69 years old from across the United Kingdom between 2006 and 2010^7^. The study received ethical approval from the National Research Ethics Service (11/NW/0382), and all participants gave written informed consent. This research has been conducted under application 40616. To construct the knowledge graph (KG), we included 4,280 individuals who had both imaging data and a documented diagnosis of AF, HF, MI, HCM, or DCM, based on ICD-9 and ICD-10 codes (Supplementary Table 1). Additionally, a reference group of 5,304 healthy participants was incorporated to ensure a broad representation of phenotypic variability. However, our primary emphasis was placed on the three most common conditions: HF, AF, and MI.

### Image-derived cardiovascular phenotypes

CMR imaging was performed on participants to capture two-dimensional retrospectively-gated cine imaging on a 1.5T magnet (Siemens Healthineers, Erlangen, Germany)^41^. Cine images were acquired in the left ventricular short-axis from base to apex, as well as in the long-axis two-and four-chamber views. Each cine series consisted of 50 cardiac frames, with a typical temporal resolution of 31 ms. Additional transverse cine images of the ascending and descending thoracic aorta were also obtained.

The two-dimensional cine images in both short- and long-axis views were segmented using fully convolutional neural networks, achieving segmentation quality comparable to that of expert human readers^8^ (Figure 3). From these segmentations, quantitative measurements of the left and right ventricles were obtained, including end-diastolic volume, end-systolic volume, stroke volume, and ejection fraction. Left ventricular mass was computed by multiplying myocardial volume by a density of 1.05 g·ml^-1^.

Atrial volumes were calculated using the biplane area-length method, where *A*_2*Ch*_ and *A*_4*Ch*_ represent the atrial areas measured in the two- and four-chamber cine views, respectively, and *L* denotes the average longitudinal diameter across both views. The formula used was: 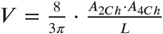

All CMR-derived measurements were indexed to body surface area (BSA), which was calculated using the Du Bois formula: BSA = 0.007184 * Height^0.725^ * Weight^0.425^ with height in cm and weight in kg.Left ventricular wall thickness was measured at end-diastole as the distance between the segmented epicardial and endocardial borders.

Circumferential and radial strains were computed from cine short-axis images using the formula 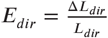 with *dir* where *dir* represents either the circumferential or radial direction, *L*_*dir*_ is the initial length of a line segment in that direction, and Δ*L*_*dir*_ denotes the change in length over time. Longitudinal strain was estimated from motion tracking in the long-axis four-chamber view. Segmentation of the aortic cine images was performed using a spatiotemporal neural network^42^. From these segmentations, the maximum and minimum cross-sectional areas were extracted. Aortic distensibility was then calculated using central blood pressure values, obtained via peripheral pulse wave analysis (Vicorder, Wuerzburg, Germany)^43^.

### Data Extraction

Data from participants in UK Biobank included CMR, disease records, air pollution exposure, medication history, age, gender, ethnicity, and body mass index. These variables represent a comprehensive dataset capturing both phenotypic and environmental factors, allowing for a holistic analysis of the relationships between genetic predispositions, lifestyle factors, and disease outcomes. CMR imaging provides detailed structural and functional cardiovascular parameters which assess cardiac volumes and mass, diastolic function, and aortic distensibility. Inclusion criteria for the KG experiments and outcome analysis are shown in Figure 2. Eighteen external databases were used in the KG spanning a variety of biomedical domains including genetic annotations, protein interactions, pathway enrichment, disease ontologies, and drug-target interactions. Details of the external databases are given below:

#### Gene Ontology

Gene Ontology (GO) is a comprehensive and widely-used resource that provides a standardised framework for representing the attributes of genes and gene products across all species^44^. Developed to unify biological terminology, it serves as a tool for organising and interpreting vast amounts of genomic and proteomics data. GO categorises biological knowledge into three main domains: molecular functions, which describe the specific activities of a gene product, such as enzyme activity or binding affinity; biological processes, which represent the broader series of events or pathways that the gene or gene product contributes to, such as cell division or signal transduction; and cellular components, which indicate the specific locations within a cell where a gene product is active, such as the nucleus, membrane, or extracellular region.

#### Human Phenotype Ontology

The Human Phenotype Ontology (HPO) provides a structured and standardised vocabulary to describe human phenotypic abnormalities, linking them to underlying genetic causes^45^. It is a resource for integrating phenotypic information across different datasets, facilitating the systematic analysis of phenotype-genotype correlations. The HPO framework is organised hierarchically capturing phenotypic abnormalities at varying levels of granularity, from broad categories, such as “abnormality of the cardiovascular system” to specific terms, such as “atrial septal defect”. Its curated dataset integrates knowledge from diverse sources, including clinical reports, literature, and existing biomedical ontologies.

#### Bgee

Bgee is a comprehensive database of gene expression data across multiple species, offering a curated and standardised resource for comparative transcriptomics and functional genomics^46^. Bgee integrates gene expression data derived from multiple experimental platforms, including RNA-seq, microarrays, and in situ hybridisation, ensuring representation of gene expression patterns across diverse biological contexts. For this study, we retrieved gene expression data for humans from Bgee to provide insights into tissue- and organ-specific gene expression profiles. Bgee employs ontologies such as the Uberon multi-species anatomy ontology and developmental stage ontologies to annotate gene expression data systematically. This facilitates cross-species comparisons to investigate the evolutionary conservation of gene expression patterns in homologous tissues and organs. The integration of these ontologies also supports the exploration of gene functions within their anatomical and developmental contexts, providing deeper insights into the roles of genes in complex biological systems.

#### Comparative Toxicogenomics Database

The Comparative Toxicogenomics Database (CTD) is a comprehensive resource that connects information about the interactions between chemicals, genes, and diseases, enabling studies on chemical-induced diseases and toxicogenomic relationships through curated and integrated data^47^. By providing manually curated information from scientific literature, the database facilitates the exploration of complex molecular mechanisms underlying the effects of environmental exposures on health. The database also supports the identification of potential therapeutic targets and biomarkers by linking chemical exposures to specific genes, pathways, and diseases. The database organises information into structured relationships, such as chemical-gene interactions, chemical-disease associations, and gene-disease associations. Furthermore, CTD includes information about chemical properties, dose-response relationships, and exposure contexts.

#### DisGeNET

DisGeNET is a comprehensive knowledge platform that integrates data on human gene-disease associations from diverse sources, including expert curation, data from the scientific literature, databases of genetic variants, and computational predictions, to support research in genomics and disease biology (version:24.4)^12^. As one of the most extensive repositories for gene-disease relationships, DisGeNET provides a unified framework to explore the genetic underpinnings of human diseases, encompassing both Mendelian and complex diseases, as well as rare conditions. The platform organises its data into a structured and standardised format, enabling integration with other resources and tools. DisGeNET’s content is derived from a wide array of sources, such as ClinVar, GWAS Catalog, UniProt, and Orphanet, and includes both experimentally validated associations and computationally inferred links. This diversity ensures a comprehensive dataset, offering insights into the genetic basis of diseases from multiple perspectives.

#### DrugBank

DrugBank (version 5.1.8) is a comprehensive and richly curated resource that integrates detailed drug data with extensive information on their mechanisms of action, interactions, targets, and associated pathways^13^. Serving as a tool for drug development, pharmacological research, and clinical applications, DrugBank integrates chemical, pharmacological, and molecular biological data. The database bridges chemistry and biology by linking drugs to relevant proteins, genes, and pathways, enabling researchers to study the complex interplay between therapeutics and biological systems. DrugBank includes information on a wide range of drug types, including small molecules, biologics, nutraceuticals, and experimental compounds. For each drug, the database provides comprehensive annotations, including chemical structures, pharmacokinetics, mechanisms of action, drug-drug interactions, and adverse effects. Additionally, it contains detailed data on drug targets, enzymes, transporters, and carriers, along with their sequences, structures, and roles in human physiology. This supports applications such as drug repurposing, biomarker discovery, and precision medicine.

#### DrugCentral

DrugCentral is a comprehensive resource that provides up-to-date, detailed information on FDA-approved drugs, including their indications, mechanisms of action, pharmacological properties, and therapeutic uses^48^. It is designed to support a wide range of pharmaceutical research and development activities, including drug discovery, repurposing, and repositioning studies. DrugCentral facilitates the identification of promising drug candidates, the exploration of new therapeutic indications, and the optimisation of existing treatments. DrugCentral includes extensive information on the chemical, pharmacokinetic, and pharmacodynamic profiles of FDA-approved drugs. For each drug, the database provides details such as chemical structure, molecular weight, drug classification, routes of administration, and therapeutic indications. Additionally, the resource offers insights into the mechanisms of action, targets, and molecular pathways involved, enabling a deeper understanding of how drugs exert their effects at the molecular and cellular levels.

#### Mayo Clinic

Information linking symptoms, causes, risk factors, complications, and prevention of 2,227 diseases and conditions was re-used from publicly available web-scraped data from the Mayo Clinic knowledgebase^4^.

#### MONDO Disease Ontology

MONDO is a unified disease ontology that integrates a wide array of disease classification systems, offering a harmonised framework for disease annotations and biomedical research^49^. By incorporating and standardising information from diverse resources, MONDO facilitates consistent and comprehensive disease categorisation, improving the interoperability and integration of data across various research fields. This integrative approach enhances the ability to link disease phenotypes to underlying genotypes, enabling a deep understanding of disease mechanisms and their genetic foundations. MONDO provides a standardised terminology for disease classification that encompasses a wide range of diseases, from rare genetic disorders to common multifactorial conditions.

#### Orphanet

Orphanet is a comprehensive reference portal dedicated to rare diseases and orphan drugs^50^. It provides an extensive collection of resources that include disease descriptions, diagnostic criteria, genetic underpinnings, epidemiology, and clinical management guidelines. The portal facilitates the identification and understanding of rare conditions, which are often overlooked due to their low prevalence. The database also includes detailed information on orphan drugs, which are critical for treating rare diseases, and offers access to regulatory information, market authorisation, and clinical usage.

#### Reactome

Reactome is an open-source pathway database that provides detailed information about the molecular mechanisms underlying biological processes^51^. It offers comprehensive insights into cellular pathways, their components, and how these pathways interconnect with various physiological and pathological states. By mapping the flow of biological information across different molecular events, Reactome enables a deep understanding of cellular processes, such as signal transduction, gene expression, metabolism, and immune response, as well as their associations with diseases. Reactome contains curated pathway annotations, which are continuously updated to reflect current research and experimental data. The database includes a wide variety of biological pathways, spanning simple molecular interactions to complex multi-step processes involving various biomolecules, such as proteins, lipids, and nucleic acids. These pathways are not only specific to humans but also cover pathways from model organisms, allowing for cross-species comparisons and a broader understanding of conserved biological mechanisms.

#### The Side Effect Resource (SIDER)

SIDER (version: 4.1) is a comprehensive database that provides structured information on the side effects of marketed drugs, offering insights into drug safety and pharmacovigilance^52^. The data is compiled from publicly available information, including package inserts, clinical trial reports, and post-marketing surveillance data. SIDER contains detailed records of side effects associated with a wide range of FDA-approved drugs, capturing both common and rare adverse events. The database includes information on the frequency, severity, and type of side effects, as well as the specific patient populations that may be more vulnerable to these adverse reactions.

#### Uberon

Uberon is an integrated, cross-species ontology that provides a comprehensive and standardised description of anatomical structures across a wide range of organisms^53^. Uberon facilitates the integration of anatomical knowledge from different species, enabling researchers to explore the similarities and differences in the structures of various organisms. This ontology bridges the gap between species, allowing for a unified understanding of biological form and function. Uberon is designed to be a multi-species resource, covering not only human anatomy but also the anatomy of model organisms (such as mice, zebrafish, and fruit flies) and non-model species, including plants and microorganisms. This broad scope allows for the comparison of anatomical structures across species, providing insights into conserved features, evolutionary adaptations, and the functional roles of specific organs and tissues.

#### STRING

STRING (version:11.5) is a comprehensive database that provides detailed information on known and predicted protein-protein interactions (PPIs), integrating an array of data sources, including experimental results, curated databases, and computational predictions^54^. The database facilitates the exploration of functional associations between proteins, providing insights into the molecular networks that drive cellular processes. By compiling data from multiple sources, STRING offers a high-confidence representation of protein interactions, enabling researchers to construct robust protein interaction networks and better understand how proteins collaborate to maintain cellular function. The STRING database integrates information from a diverse range of experimental methods, including yeast two-hybrid screens, affinity purification followed by mass spectrometry (AP-MS), co-immunoprecipitation, and other high-throughput techniques. Additionally, STRING incorporates data from curated databases, such as those focused on biochemical pathways and molecular function, as well as computational predictions based on sequence homology, text mining, and structural modeling.

#### BioGRID

BioGRID (version:4.4.198) is a comprehensive, curated repository that provides high-quality data on protein, genetic, and chemical interactions across a wide range of organisms, including humans, model organisms, and pathogens^55^. By consolidating diverse interaction data from various sources, BioGRID plays a role in supporting molecular biology, systems biology, and biomedical research. The database is designed to facilitate the exploration of molecular networks, offering insights into how proteins, genes, and chemicals interact to regulate cellular processes, maintain homeostasis, and contribute to disease mechanisms. BioGRID curates data from a variety of experimental techniques, including yeast two-hybrid screens, co-immunoprecipitation, affinity purification-mass spectrometry (AP-MS), and synthetic lethal screens. In addition to experimental data, BioGRID integrates information from computational predictions, enabling researchers to investigate both direct and indirect interactions within complex biological systems. This multi-source approach ensures that the data within BioGRID is comprehensive, covering a wide range of biological interactions that help researchers gain a deeper understanding of cellular functions. The protein interaction data in BioGRID is valuable for constructing molecular interaction networks.

#### Gene Regulation

Gene regulation databases play a critical role in understanding the complex mechanisms that control gene expression, including the processes that regulate the transcriptional, post-transcriptional, and epigenetic aspects of gene activity (version:2.0)^56^. These databases integrate a wide range of information about how genes are turned on or off in response to various signals and how they maintain their expression patterns across different biological contexts. Specifically, these databases provide detailed insights into transcription factor binding, epigenetic modifications, non-coding RNA regulation, and other molecular interactions that govern gene expression. One of the key components of gene regulation is the binding of transcription factors (TFs) to specific DNA sequences within promoter regions or enhancers of target genes. Gene regulation databases, such as TRANSFAC and its module TRANSCompel, provide curated and experimentally validated data on TF binding sites, offering researchers the ability to predict which transcription factors regulate specific genes.

#### CORUM

CORUM (version:3.0) is a comprehensive, curated resource dedicated to cataloging protein complexes in mammals, offering insights into the composition, function, and interactions of these complexes that are essential for various cellular processes^57^. This extensive database serves as a key resource for understanding the molecular machinery underlying fundamental biological processes, including signal transduction, gene expression regulation, metabolism, and cellular structure. By integrating experimental data from various sources, CORUM provides an up-to-date repository of information on the assembly, function, and interactions of mammalian protein complexes. Protein complexes are groups of two or more proteins that interact to perform specific biological functions within the cell. The formation and regulation of these complexes are critical for maintaining cellular homeostasis, controlling signal transduction pathways, and enabling cellular responses to internal and external stimuli.

#### National Library of Medicine

The National Library of Medicine (NLM) hosts a range of biomedical and health-related resources, including the National Center for Biotechnology Information (NCBI). For the construction of the KG we incorporated gene and Gene Ontology (GO) term associations derived from the NCBI Gene database (formerly known as Entrez Gene). Specifically, we used a processed dataset, which was based on the publicly available curated associations between genes and GO terms describing biological processes, molecular functions, and cellular components^4^. The associations (n = 297,917) between gene and GO terms from this dataset were incorporated as links in the KG.

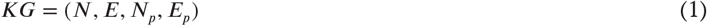

### Knowledge graph construction

Diverse biological databases were integrated to construct CardioKG. The data from various databases for each entity were harmonised by creating a dictionary that has all the possible ontologies of each node and then natural laguage processing library “nltk” was used to tokenise, clean and normalise the ontologies prior to matching with a reference list. For instance, the disease entity heart failure has various classifications such as congestive heart disease and left ventricular failure which required harmonisation. The nodes in the constructed KG were extracted from 12 different entities including genes, diseases, medication, individuals, CMR, phenotypes, anatomy, BP, CC, MF, pathways and air pollutants, along with their relationships.The CMR features were incorporated into the KG as distinct nodes, by connecting them to heart anatomical regions such as left and right ventricle (Supplementary Figure 3), with their corresponding values assigned as properties on the edges connecting the CMR nodes to other related nodes. Additionally, individuals’ sex, age, body mass index (BMI), ethnicity, and BSA were integrated as properties within the corresponding individual nodes. Hence, the final graph has nodes (*N*), properties of nodes (*N*_*p*_), edges (*E*) and properties of edges (*E*_*p*_) as shown in Equation 1:

### Knowledge graph embeddings

Graph embedding techniques aim to learn low-dimensional representations of nodes and edges in a graph while preserving the graph’s structural properties and node relationships. Graph embeddings can be used for various downstream tasks such as node clustering and link prediction. KG embedding methods, such as Node2Vec^15^ and ComplEx^17^, focus only on graph connectivity and fail to capture node and relation attributes. Therefore, DVGAE was employed as the embedding algorithm, as it integrates both node and relation properties for generating embeddings. The DVGAE is a deep learning model that uses a variational graph auto-encoder^9^. However, our DVGAE networks extend traditional VGAE by handling edge direction, node type as well as node and relation properties in the KG (supplementary Figure 19). The DVGAE has the following components:

#### Encoder

The encoder uses multiple graph convolutional network (GCN) layers to process four inputs including node type, node properties, edge properties and edge direction to learn a low-dimensional latent representation. These embeddings are further enriched with node-type information to better represent the diversity of node roles in the graph. By learning these latent variables, the encoder enables the model to effectively capture the underlying relationships in the graph required for graph reconstruction and link prediction. The latent layer contains the low-dimensional embeddings generated by the encoder component of the directed variational graph auto-encoder, which captures the underlying structural and semantic information from the directed KG. These embeddings serve as compact representations of the nodes, preserving key relational patterns, and are used as input for downstream tasks.

#### Decoder

After the encoder generates the embeddings, the decoder takes these embeddings and predicts the presence or absence of edges between nodes. Specifically, the decoder calculates the dot product between pairs of node embeddings, which gives a measure of how strongly connected two nodes are. The final output is a set of probabilities indicating whether there should be an edge between each pair of nodes. The decoder thus enables the model to reconstruct the graph structure based on the learned node embeddings.

#### Validation

For graph-based learning tasks, it is essential for the model to capture the global structure of the graph, including relationships between distant nodes. To train the DVGAE model, the dataset was split into training (70%) and validation (30%) subsets. The embeddings produced by the encoder are used to compute the reconstruction loss and the Kullback–Leibler (KL) divergence loss, as defined in Equations 2 and 3. The reconstruction loss evaluates the model’s ability to correctly predict edges, while the KL loss regularizes the latent space during training. Training is carried out using a grid search approach over a set of hyperparameters including learning rate {0.001, 0.01, 0.1}, number of epochs {50, 100, 200}, and latent dimension sizes {25, 50, 100}. For each combination, the model is trained and evaluated on the validation set. The optimal configuration—learning rate of 0.001, 100 epochs, and a latent dimension size of 50—was selected based on the lowest loss. Validation loss is computed after each epoch by comparing the predicted edges to the true edges in the validation set. Negative edges are randomly sampled from node pairs that do not exist in the original graph, providing a contrastive signal for learning. The validation loss, defined in Equation 4, serves as the basis for selecting the final model, with the configuration yielding the lowest validation loss chosen as optimal.

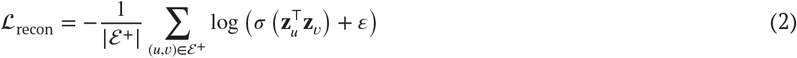

where ℰ^+^ denotes the set of positive (observed) edges, **z**_u_ and **z**_v_ are the learned embedding vectors for nodes u and υ,σ(·) is the sigmoid function, and ε is a small constant added for numerical stability.

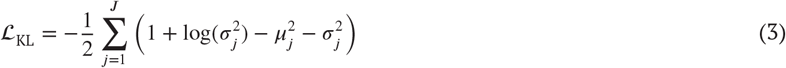

where *μ*_*j*_ and σ_*j*_ represent the *j*-th dimension of the mean and standard deviation vectors output by the encoder for a given node, and ***J*** is the size of the latent space

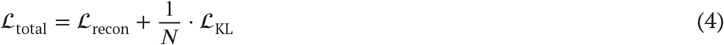

The final DVGAE model, configured with the optimized hyperparameters, was used to generate node embeddings. To evaluate the quality of these embeddings, edge-wise accuracy (Equation 5) was employed as an evaluation metric. By comparing the reconstructed graph to the original graph on an edge-by-edge basis, this metric provides a quantitative assessment of how well the learned embeddings preserve the structural properties of the input graph.

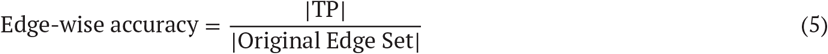

Where True Positives (TP) is the intersection of the original and reconstructed edge sets.

### Predicting novel gene-disease associations

#### Heart failure

The embedding vector of the “heart failure” node was concatenated with the embedding vectors of both positive and negative genes (Supplementary Table 3). Positive genes refer to those with a known association with HF, as identified from the standard database DisGeNET. Although DisGeNET was used in constructing the knowledge graph, only phenotype-gene and other non-evaluated associations were included, ensuring that no data leakage occurred during the predictive task. Negative genes, on the other hand, are those with no association with cardiovascular diseases, including HF, according to DisGeNET. The concatenated embeddings were assigned labels of either 0 or 1, as defined in Equation 6 below.

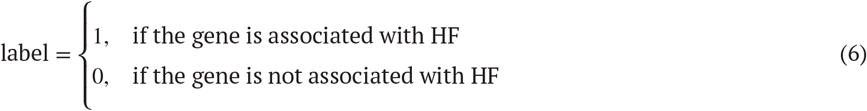

Subsequently, three machine learning (ML) algorithms—random forest (RF, hyperparameters (n-trees, max-features, max-depth, min-samples-split and min-samples-leaf)), support vector machine (SVM, hyperparameters (C, gamma and kernel)) and artificial neural networks (ANNs, hyperparameters (learning rate, dropout rate, batch size and epochs))—were trained on the concatenated embeddings to predict the association between the genes and disease using 5-fold cross-validation. Each model was optimized using relevant hyperparameters: RF (number of trees, maximum features, maximum depth, minimum samples for split, and minimum samples per leaf), SVM (regularization parameter C, kernel coefficient gamma, and kernel type), and ANN (learning rate, dropout rate, batch size, and number of epochs). The performance of the classifiers was evaluated based on accuracy (as defined in Equation 7), Area Under the Receiver Operating Characteristic Curve (AUCROC), sensitivity and specificity. The classifier demonstrating the best performance was then selected and used to predict new associations between heart failure and genes that have unknown associations with HF. Each gene predicted to have a positive association with HF was assigned a probability by the machine learning model, which was then used to rank the predicted genes.

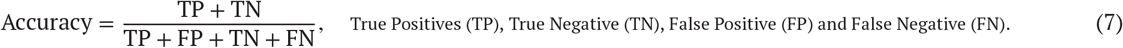

#### Atrial fibrillation

The embedding of “atrial fibrillation” node was concatenated with the embeddings of both positive and negative genes (Supplementary Table 7). Positive genes refer to those known to be associated with AF, while negative genes have no association with cardiovascular diseases, including AF. The same approach used for heart failure was then applied to AF to predict new associations.

#### Myocardial infarction

In MI, embedding of “myocardial infarction” node was concatenated with the embeddings of both positive and negative genes (Supplementary Table 5). Then, the same approach used for HF and AF was applied to predict new associations.

### Enrichment analysis

The analysis was performed for each disease separately to validate the predicted genes. In each disease, the top ten genes, ranked by their predicted probabilities from the ML classifier, were subjected to enrichment analysis. This analysis aimed to identify whether these predicted genes were associated with critical pathways relevant to each disease, including AF, HF and MI. The `g:Profiler`platform https://biit.cs.ut.ee/gprofiler/gost was utilised to perform the enrichment analysis, identifying significantly enriched pathways associated with the top ten predicted genes in each disease. The *P* values of the enriched pathways were adjusted using the Benjamini-Hochberg method to control for multiple testing. Subsequently, the enriched pathways were examined to determine whether they included relevant pathways associated with the disease. Hence, genes associated with pathways identified as critical for the disease were considered to be associated with the disease.

### Druggablity analysis

The druggability of the top ten predicted genes for each disease was evaluated using the DGIdb^14^. This analysis aimed to identify which of the predicted genes protein could be effectively targeted and modulated by existing medications, providing insights into their potential as therapeutic targets. Five types of drug–gene interactions were considered in this analysis: agonist, inhibitor, modulator, activator, and antibody. These were selected because they represent the most common and therapeutically relevant interaction types associated with druggable genes in prior studies and databases such as DGIdb^58^. While DGIdb includes additional interaction types (e.g., binder), our focus was on those with clearer functional implications in gene regulation and therapeutic modulation.

### Assessing the significance of imaging data

The significance of the CMR features was assessed from two distinct perspectives including the structural characteristics of the graph and the influence on the predicted gene associations:

#### Structural characteristics of the graph

The CMR features were integrated as nodes in the KG and to understand how influential they are the PageRank algorithm^22^ was applied to calculate scores for each node in the KG. PageRank is a centrality algorithm identifies highly influential nodes within the graph. It considers the entire structure of the graph, incorporating both direct and indirect relationships to quantify a node’s influence as a score. In biological networks, such as gene-disease association graphs, PageRank can highlight critical nodes that influence overall connectivity and information flow.

#### Influence on the predicted genes

To assess the influence of CMR features on the predicted gene-disease associations, the CMR nodes were excluded from the KG. The same methodology was then applied, including generating node embeddings, predicting gene-disease associations, and conducting enrichment analysis. The results were compared to the scenario that included the CMR nodes to determine whether the predicted genes differed. Additionally, the analysis examined whether the inclusion of CMR features led to the prediction of a greater number of pathways associated with the diseases of interest (AF, HF and MI). The number of identified critical pathways for each disease was calculated and employed to quantify the differences in predicted pathways between the two scenarios, then a Chi-squared test was conducted to evaluate the difference between the two scenarios in terms of the number of identified pathways associated with the diseases.

### Drug repurposing

In drug repurposing, the same cohort was used (n = 9,584) and the embeddings representing the disease of interest were concatenated with the embeddings of medications that are either indicated or contraindicated in that disease as defined in Equation 8 (Supplementary Tables 15 to 17). We used a ground truth of known medication–disease pairs from DrugBank^13^ database. Although DrugBank was incorporated in building the knowledge graph, only other non-assessed associations were included to prevent any data leakage during the predictive task. Three machine learning algorithms—support vector machines (SVM), random forests (RF), and artificial neural networks (ANN)—were then trained and validated using this integrated data to predict novel associations between the disease of interest and previously un-associated medications. The validation of the top ten predicted medications (based on their assigned probabilities) was conducted in two phases. First, an enrichment analysis was performed on the drug targets to determine whether they were significantly associated with critical pathways related to the disease of interest. In the second, survival analysis was carried out to assess the survival probability of individuals (with death as the outcome) with the disease who received the predicted medication. These individuals (n=489) represent a subset of patients who were not part of the primary cohort of 9,584 and do not have imaging phenotype data, as such data were not required for this validation step. The survival functions were adjusted for age, sex, BMI and ethnicity. Additionally, a graph-based validation was conducted by computing the cosine similarity (see Equation 9) between the predicted medication and those currently used for the treatment or management of heart failure.

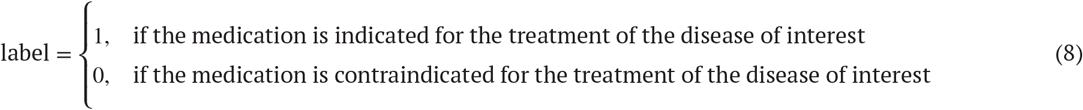

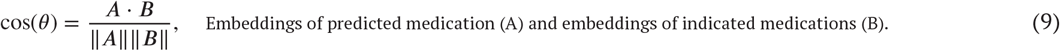

### Statistics

All analyses were performed using Python `(version 3.12.1)`and Neo4j Desktop `(version 5.20.0)`. Categorical variables were expressed as frequencies and percentages, while continuous variables were reported as medians with interquartile ranges (IQR). In `g:Profiler`, the hypergeometric test was used to calculate the *P* values for assessing the significance of the association between genes and pathways. Subsequently, Benjamini-Hochberg method was applied to control the false discovery rate by adjusting the *P* values for multiple comparisons and a *P* value < 0.05 was considered statistically significant. The chi-squared test was performed to assess the importance of the CMR features in terms of the number of identified pathways associated with the diseases. The survival functions were estimated based on fitted Cox models.

## Funding

The study was supported by the Medical Research Council (MC_UP_1605/13); the British Heart Foundation (RG/F/24/110138, RE/24/130023, CH/F/24/90015, FS/IPBSRF/22/27059); Bayer AG, Sir Jules Thorn Charitable Trust [21JTA], and the National Institute for Health Research (NIHR) Imperial College Biomedical Research Centre. D.P.O’R. and J.S.W. are supported by the British Heart Foundation’s Big Beat Challenge award to CureHeart (BBC/F/21/220106).

## Disclosures

D.P.O’R. receives research support from Bayer AG and Calico Labs, and is a paid consultant to Bayer AG and Bristol Myers Squibb. J.S.W. has received research support from Bristol Myers Squibb, has acted as a paid advisor to Health Lumen, Tenaya Therapeutics, and Solid Biosciences, and is a founder with equity in Saturnus Bio.

## Author contributions

Conceptualization: D.P.O’R., K.R.; Methodology: K.R., D.P.O’R., C.B., K.McG., L.C., S.Z., S.K., S.T., P.G., J.Z.; Formal analysis:K.R., C.B.; Resources: D.P.O’R., W.B.; Software: D.S., V.L., K.R.; Writing - original draft: K.R.; Writing - review & editing: K.R.,D.P.O’R., A.de M., J.W., L.C., C.B., L.Z., M.I., V.K.; Visualization: K.R., D.P.O’R.; Supervision: D.P.O’R.; Project administration: D.P.O’R.; Funding acquisition: D.P.O’R.

## Data availability

The scripts for data analysis are publicly available at https://github.com/ImperialCollegeLondon/cardioKG (DOI:10.5281/zenodo.16025952). Data from UK Biobank are available for approved research.

## Notes

### Author Declarations

The study received ethical approval from the National Research Ethics Service (11/NW/0382), and all participants gave written informed consent.

## References

1. Visscher PM, Wray NR, Zhang Q, Sklar P, McCarthy MI, Brown MA, and Yang J. 10 Years of GWAS Discovery: Biology, Function, and Translation. Am J Hum Genet. 2017;101:5–22. DOI: 10.1016/j.ajhg.2017.06.005.

2. Callahan TJ, Tripodi IJ, Stefanski AL, Cappelletti L, Taneja SB, Wyrwa JM, Casiraghi E, Matentzoglu NA, Reese J, Silverstein JC, et al. An open source knowledge graph ecosystem for the life sciences. Sci Data. 2024;11:363. DOI: 10.1038/s41597-024-03171-w.

3. Li L, Wang P, Yan J, Wang Y, Li S, Jiang J, Sun Z, Tang B, Chang TH, Wang S, et al. Real-world data medical knowledge graph: construction and applications. Artif Intell Med. 2020;103:101817. DOI: 10.1016/j.artmed.2020.101817.

4. Chandak P, Huang K, and Zitnik M. Building a knowledge graph to enable precision medicine. Sci Data. 2023;10:67. DOI: 10.1038/s41597-023-01960-3.

5. Ji S, Pan S, Cambria E, Marttinen P, and Yu PS. A Survey on Knowledge Graphs: Representation, Acquisition, and Applications. IEEE Trans Neural Netw Learn Syst. 2022;33:494–514. DOI: 10.1109/tnnls.2021.3070843.

6. Gualdi F, Oliva B, and Pinero J. Predicting gene disease associations with knowledge graph embeddings for diseases with curtailed information. NAR Genom Bioinform. 2024;6:lqae049. DOI: 10.1093/nargab/lqae049.

7. Bycroft C, Freeman C, Petkova D, Band G, Elliott LT, Sharp K, Motyer A, Vukcevic D, Delaneau O, O’Connell J, et al. The UK Biobank resource with deep phenotyping and genomic data. Nature. 2018;562:203–9. DOI: 10.1038/s41586-018-0579-z.

8. Bai W, Sinclair M, Tarroni G, Oktay O, Rajchl M, Vaillant G, Lee AM, Aung N, Lukaschuk E, Sanghvi MM, et al. Automated cardiovascular magnetic resonance image analysis with fully convolutional networks. J Cardiovasc Magn Reson. 2018;20:65. DOI: 10.1186/s12968-018-0471-x.

9. Lei L, Han K, Wang Z, Shi C, Wang Z, Dai R, Zhang Z, Wang M, and Guo Q. Attention-guided variational graph autoencoders reveal heterogeneity in spatial transcriptomics. Brief Bioinform. 2024;25. DOI: 10.1093/bib/bbae173.

10. Xie L, Peng YQ, and Shen X. Identifying therapeutic target genes for diabetic retinopathy using systematic druggable genome-wide Mendelian randomization. Diabetol Metab Syndr. 2025;17:145. DOI: 10.1186/s13098-025-01710-y.

11. Xu H, Aldrich MC, Chen Q, Liu H, Peterson NB, Dai Q, Levy M, Shah A, Han X, Ruan X, et al. Validating drug repurposing signals using electronic health records: a case study of metformin associated with reduced cancer mortality. J Am Med Inform Assoc. 2015;22:179–91. DOI: 10.1136/amiajnl-2014-002649.

12. Pinero J, Ramirez-Anguita JM, Sauch-Pitarch J, Ronzano F, Centeno E, Sanz F, and Furlong LI. The DisGeNET knowledge platform for disease genomics: 2019 update. Nucleic Acids Res. 2020;48:D845–D855. DOI: 10.1093/nar/gkz1021.

13. Wishart DS, Feunang YD, Guo AC, Lo EJ, Marcu A, Grant JR, Sajed T, Johnson D, Li C, Sayeeda Z, et al. DrugBank 5.0: a major update to the DrugBank database for 2018. Nucleic Acids Res. 2018;46:D1074–D1082. DOI: 10.1093/nar/gkx1037.

14. Freshour SL, Kiwala S, Cotto KC, Coffman AC, McMichael JF, Song JJ, Griffith M, Griffith OL, and Wagner AH. Integration of the Drug-Gene Interaction Database (DGIdb 4.0) with open crowdsource efforts. Nucleic Acids Res. 2021;49:D1144– D1151. DOI: 10.1093/nar/gkaa1084.

15. Grover A and Leskovec J. node2vec: Scalable Feature Learning for Networks. KDD. 2016;2016:855–64. DOI: 10.1145/2939672.2939754.

16. Bordes A, Usunier N, Garcia-Durán A, Weston J, and Yakhnenko O. Translating embeddings for modeling multi-relational data. In: Proceedings of the 27th International Conference on Neural Information Processing Systems - Volume 2. NIPS’13. Lake Tahoe, Nevada: Curran Associates Inc., 2013:2787–95.

17. Trouillon T, Welbl J, Riedel S, Gaussier É, and Bouchard G. Complex embeddings for simple link prediction. In: Proceedings of the 33rd International Conference on International Conference on Machine Learning - Volume 48. ICML’16. New York, NY, USA: JMLR.org, 2016:2071–80.

18. Zheng SL, Henry A, Cannie D, Lee M, Miller D, McGurk KA, Bond I, Xu X, Issa H, Francis C, et al. Genome-wide association analysis provides insights into the molecular etiology of dilated cardiomyopathy. Nat Genet. 2024;56:2646– 58. DOI: 10.1038/s41588-024-01952-y.

19. Floyd JS, Sitlani CM, Doyle MF, Feinstein MJ, Olson NC, Heckbert SR, Huber SA, Tracy RP, Psaty BM, and Delaney JAC. Immune cell subpopulations as risk factors for atrial fibrillation: The Cardiovascular Health Study and Multi-Ethnic Study of Atherosclerosis. Heart Rhythm. 2023;20:315–7. DOI: 10.1016/j.hrthm.2022.10.012.

20. Zhai Y, Yang J, Zhang J, Yang J, Li Q, and Zheng T. Src-family Protein Tyrosine Kinases: A promising target for treating Cardiovascular Diseases. Int J Med Sci. 2021;18:1216–24. DOI: 10.7150/ijms.49241.

21. Chang CY, Yeh YH, Chan YH, Liu JR, Chang SH, Lee HF, Wu LS, Yen KC, Kuo CT, and See LC. Dipeptidyl peptidase-4 inhibitor decreases the risk of atrial fibrillation in patients with type 2 diabetes: a nationwide cohort study in Taiwan. Cardiovasc Diabetol. 2017;16:159. DOI: 10.1186/s12933-017-0640-5.

22. Johannes M, Brase JC, Fröhlich H, Gade S, Gehrmann M, Fälth M, Sültmann H, and Beißbarth T. Integration of pathway knowledge into a reweighted recursive feature elimination approach for risk stratification of cancer patients. Bioinformatics. 2010;26:2136–44. DOI: 10.1093/bioinformatics/btq345.

23. Munzel T, Camici GG, Maack C, Bonetti NR, Fuster V, and Kovacic JC. Impact of Oxidative Stress on the Heart and Vasculature: Part 2 of a 3-Part Series. J Am Coll Cardiol. 2017;70:212–29. DOI: 10.1016/j.jacc.2017.05.035.

24. Xie W, Santulli G, Reiken SR, Yuan Q, Osborne BW, Chen BX, and Marks AR. Mitochondrial oxidative stress promotes atrial fibrillation. Sci Rep. 2015;5:11427. DOI: 10.1038/srep11427.

25. Pfenniger A, Yoo S, and Arora R. Oxidative stress and atrial fibrillation. J Mol Cell Cardiol. 2024;196:141–51. DOI:10.1016/j.yjmcc.2024.09.011.

26. Roth GA, Mensah GA, Johnson CO, Addolorato G, Ammirati E, Baddour LM, Barengo NC, Beaton AZ, Benjamin EJ, Benziger CP, et al. Global Burden of Cardiovascular Diseases and Risk Factors, 1990-2019: Update From the GBD 2019 Study. J Am Coll Cardiol. 2020;76:2982–3021. DOI: 10.1016/j.jacc.2020.11.010.

27. Omidiran O, Patel A, Usman S, Mhatre I, Abdelhalim H, DeGroat W, Narayanan R, Singh K, Mendhe D, and Ahmed Z. GWAS advancements to investigate disease associations and biological mechanisms. Clin Transl Discov. 2024;4. DOI: 10.1002/ctd2.296.

28. Gusev A, Ko A, Shi H, Bhatia G, Chung W, Penninx BW, Jansen R, de Geus EJ, Boomsma DI, Wright FA, et al. Integrative approaches for large-scale transcriptome-wide association studies. Nat Genet. 2016;48:245–52. DOI: 10.1038/ng.3506.

29. Fang C, Arango Argoty GA, Kagiampakis I, Khalid MH, Jacob E, Bulusu KC, and Markuzon N. Integrating knowledge graphs into machine learning models for survival prediction and biomarker discovery in patients with non-small-cell lung cancer. J Transl Med. 2024;22:726. DOI: 10.1186/s12967-024-05509-9.

30. Nunes S, Sousa RT, and Pesquita C. Multi-domain knowledge graph embeddings for gene-disease association prediction.J Biomed Semantics. 2023;14:11. DOI: 10.1186/s13326-023-00291-x.

31. Renaux A, Terwagne C, Cochez M, Tiddi I, Nowe A, and Lenaerts T. A knowledge graph approach to predict and interpret disease-causing gene interactions. BMC Bioinformatics. 2023;24:324. DOI: 10.1186/s12859-023-05451-5.

32. Sanna GD, Nusdeo G, Piras MR, Forteleoni A, Murru MR, Saba PS, Dore S, Sotgiu G, Parodi G, and Ganau A. Cardiac Abnormalities in Alzheimer Disease: Clinical Relevance Beyond Pathophysiological Rationale and Instrumental Findings? JACC Heart Fail. 2019;7:121–8. DOI: 10.1016/j.jchf.2018.10.022.

33. Landen JW, Andreasen N, Cronenberger CL, Schwartz PF, Borjesson-Hanson A, Ostlund H, Sattler CA, Binneman B, and Bednar MM. Ponezumab in mild-to-moderate Alzheimer’s disease: Randomized phase II PET-PIB study. Alzheimers Dement (N Y). 2017;3:393–401. DOI: 10.1016/j.trci.2017.05.003.

34. Salloway S, Marshall GA, Lu M, and Brashear HR. Long-Term Safety and Efficacy of Bapineuzumab in Patients with Mild-to-Moderate Alzheimer’s Disease: A Phase 2, Open-Label Extension Study. Curr Alzheimer Res. 2018;15:1231–43. DOI: 10.2174/1567205015666180821114813.

35. Mangoni AA, Wiese MD, Woodman RJ, Sotgia S, Zinellu A, Carru C, Hulin JA, Shanahan EM, and Tommasi S. Methotrexate, blood pressure and arterial function in rheumatoid arthritis: study protocol. Future Cardiol. 2024;20:671–83. DOI: 10.1080/14796678.2024.2411167.

36. Mangoni AA, Tommasi S, Zinellu A, Sotgia S, Carru C, Piga M, and Erre GL. Repurposing existing drugs for cardiovascular risk management: a focus on methotrexate. Drugs Context. 2018;7:212557. DOI: 10.7573/dic.212557.

37. Johnson TM, Sayles HR, Baker JF, George MD, Roul P, Zheng C, Sauer B, Liao KP, Anderson DR, Mikuls TR, et al. Investigating changes in disease activity as a mediator of cardiovascular risk reduction with methotrexate use in rheumatoid arthritis. Ann Rheum Dis. 2021;80:1385–92. DOI: 10.1136/annrheumdis-2021-220125.

38. Yang J, He J, Ismail M, Tweeten S, Zeng F, Gao L, Ballinger S, Young M, Prabhu SD, Rowe GC, et al. HDAC inhibition induces autophagy and mitochondrial biogenesis to maintain mitochondrial homeostasis during cardiac ischemia/reperfusion injury. J Mol Cell Cardiol. 2019;130:36–48. DOI: 10.1016/j.yjmcc.2019.03.008.

39. Peng C, Xia F, Naseriparsa M, and Osborne F. Knowledge Graphs: Opportunities and Challenges. Artif Intell Rev. 2023:1–32. DOI: 10.1007/s10462-023-10465-9.

40. Hou Y, Yeung J, Xu H, Su C, Wang F, and Zhang R. From Answers to Insights: Unveiling the Strengths and Limitations of ChatGPT and Biomedical Knowledge Graphs. Res Sq. 2023. DOI: 10.21203/rs.3.rs-3185632/v1.

41. Petersen SE, Matthews PM, Francis JM, Robson MD, Zemrak F, Boubertakh R, Young AA, Hudson S, Weale P, Garratt S, et al. UK Biobank’s cardiovascular magnetic resonance protocol. J Cardiovasc Magn Reson. 2016;18:8. DOI: 10.1186/s12968-016-0227-4.

42. Bai W, Suzuki H, Qin C, Tarroni G, Oktay O, Matthews PM, and Rueckert D. Recurrent Neural Networks for Aortic Image Sequence Segmentation with Sparse Annotations. Medical Image Computing and Computer Assisted Intervention – MICCAI 2018. 2018:586–94. DOI: 10.1007/978-3-030-00937-3_67.

43. Bai W, Suzuki H, Huang J, Francis C, Wang S, Tarroni G, Guitton F, Aung N, Fung K, Petersen SE, et al. A population-based phenome-wide association study of cardiac and aortic structure and function. Nat Med. 2020;26:1654–62. DOI: 10.1038/s41591-020-1009-y.

44. Gene Ontology C. The Gene Ontology resource: enriching a GOld mine. Nucleic Acids Res. 2021;49:D325–D334. DOI:10.1093/nar/gkaa1113.

45. Kohler S, Vasilevsky NA, Engelstad M, Foster E, McMurry J, Ayme S, Baynam G, Bello SM, Boerkoel CF, Boycott KM, et al. The Human Phenotype Ontology in 2017. Nucleic Acids Res. 2017;45:D865–D876. DOI: 10.1093/nar/gkw1039.

46. Bastian FB, Roux J, Niknejad A, Comte A, Fonseca Costa SS, de Farias TM, Moretti S, Parmentier G, de Laval VR, Rosikiewicz M, et al. The Bgee suite: integrated curated expression atlas and comparative transcriptomics in animals. Nucleic Acids Res. 2021;49:D831–D847. DOI: 10.1093/nar/gkaa793.

47. Davis AP, Grondin CJ, Johnson RJ, Sciaky D, Wiegers J, Wiegers TC, and Mattingly CJ. Comparative Toxicogenomics Database (CTD): update 2021. Nucleic Acids Res. 2021;49:D1138–D1143. DOI: 10.1093/nar/gkaa891.

48. Avram S, Bologa CG, Holmes J, Bocci G, Wilson TB, Nguyen DT, Curpan R, Halip L, Bora A, Yang JJ, et al. DrugCentral 2021 supports drug discovery and repositioning. Nucleic Acids Res. 2021;49:D1160–D1169. DOI: 10.1093/nar/gkaa997.

49. Shefchek KA, Harris NL, Gargano M, Matentzoglu N, Unni D, Brush M, Keith D, Conlin T, Vasilevsky N, Zhang XA, et al. The Monarch Initiative in 2019: an integrative data and analytic platform connecting phenotypes to genotypes across species. Nucleic Acids Res. 2020;48:D704–D715. DOI: 10.1093/nar/gkz997.

50. Weinreich SS, Mangon R, Sikkens JJ, Teeuw ME, and Cornel MC. [Orphanet: a European database for rare diseases].Ned Tijdschr Geneeskd. 2008;152:518–9.

51. Jassal B, Matthews L, Viteri G, Gong C, Lorente P, Fabregat A, Sidiropoulos K, Cook J, Gillespie M, Haw R, et al. The reactome pathway knowledgebase. Nucleic Acids Res. 2020;48:D498–D503. DOI: 10.1093/nar/gkz1031.

52. Kuhn M, Letunic I, Jensen LJ, and Bork P. The SIDER database of drugs and side effects. Nucleic Acids Res. 2016;44:D1075– 9. DOI: 10.1093/nar/gkv1075.

53. Mungall CJ, Torniai C, Gkoutos GV, Lewis SE, and Haendel MA. Uberon, an integrative multi-species anatomy ontology. Genome Biol. 2012;13:R5. DOI: 10.1186/gb-2012-13-1-r5.

54. Szklarczyk D, Gable AL, Nastou KC, Lyon D, Kirsch R, Pyysalo S, Doncheva NT, Legeay M, Fang T, Bork P, et al. The STRING database in 2021: customizable protein-protein networks, and functional characterization of user-uploaded gene/measurement sets. Nucleic Acids Res. 2021;49:D605–D612. DOI: 10.1093/nar/gkaa1074.

55. Oughtred R, Rust J, Chang C, Breitkreutz BJ, Stark C, Willems A, Boucher L, Leung G, Kolas N, Zhang F, et al. The BioGRID database: A comprehensive biomedical resource of curated protein, genetic, and chemical interactions. Protein Sci. 2021;30:187–200. DOI: 10.1002/pro.3978.

56. Matys V, Kel-Margoulis OV, Fricke E, Liebich I, Land S, Barre-Dirrie A, Reuter I, Chekmenev D, Krull M, Hornischer K, et al. TRANSFAC and its module TRANSCompel: transcriptional gene regulation in eukaryotes. Nucleic Acids Res. 2006;34:D108–10. DOI: 10.1093/nar/gkj143.

57. Giurgiu M, Reinhard J, Brauner B, Dunger-Kaltenbach I, Fobo G, Frishman G, Montrone C, and Ruepp A. CORUM: the comprehensive resource of mammalian protein complexes-2019. Nucleic Acids Res. 2019;47:D559–D563. DOI: 10.1093/nar/gky973.

58. Griffith M, Griffith OL, Coffman AC, Weible JV, McMichael JF, Spies NC, Koval J, Das I, Callaway MB, Eldred JM, et al. DGIdb: mining the druggable genome. Nat Methods. 2013;10:1209–10. DOI: 10.1038/nmeth.2689.

