## Supplementary material for "A multi-modal vision knowledge graph of cardiovascular disease"

**Supplementary Tables 1 - 17**
**Supplementary Figures 1 - 19**
**Supplementary Data File 1**: `gProfiler_HF_CMR.csv`
**Supplementary Data File 2**: `gProfiler_AF_CMR.csv`
**Supplementary Data File 3**: `gProfiler_MI_CMR.csv`

**Supplementary Table 1. The used ICD-10 and ICD-9 codes to extract the disease.** The codes that have been used to extract the individuals form the UKBiobank. ICD-10 is the tenth revision of the International Classification of Diseases, a system used by healthcare providers to code and classify diseases and health conditions. It replaced ICD-9.

| Disease | ICD-10 code | ICD-9 code |
|---|---|---|
| Heart failure | I11.0, I13.0, I13.2, I50 | 428 |
| Atrial fibrillation | I48.0, I48.1, I48.2, I48.3, I48.4, I48.9 | 4273 |
| Myocardial infarction | I21, I22, I23 | 4109, 4129 410, 411, 412 |
| Hypertrophic cardiomyopathy | I42.1, I42.2 | 4251 |
| Dilated cardiomyopathy | I42.0, I42.6, I42.7 | 4254, 4255 |

**Supplementary Table 2. The demographics and the imaging characteristics of the extracted cohort from the UK Biobank.** All the individuals (n=9,584) have cardiac magnetic resonance imaging (CMR). Healthy includes individuals who have non of the following diseases: cardiovascular diseases, heart diseases, peripheral vascular disease, cerebrovascular diseases, hypertension, hyper-cholesterol, diabetes, cancer and chronic kidney disease. AF: atrial fibrillation, MI: myocardial infarction, HF: heart failure, HCM: hypertrophic cardiomyopathy, DCM: dilated cardiomyopathy, body mass index (BMI), body surface area (BSA).

| **Baseline characteristics** | **Healthy** (n=5,304) | **AF** (n=2,153) | **MI** (n=1,414) | **HF** (n=602) | **HCM** (n=56) | **DCM** (n=55) |
|---|---|---|---|---|---|---|
| Sex Males(%) | 1747(32.93%) | 1494(69.39%) | 1123 (79.42%) | 441 (73.26%) | 43 (76.79%) | 45 (81.82%) |
| Ethnicity White(%) | 4650 (87.67%) | 2107 (97.86%) | 1361 (96.25%) | 588 (97.67%) | 55 (98.21%) | 54 (98.18%) |
| Age at time of MRI (years) Median[Q1;Q3] | 61.0 [55.25; 67.0] | 70.0 [65.0; 74.0] | 69.0 [64.0; 73.0] | 71.0 [66.0; 74.0] | 67.0 [62.75; 71.0] | 69.0 [63.0; 72.0] |
| Body Mass Index (kg/m$^2$) Median[Q1;Q3] | 23.67 [21.83; 25.71] | 26.31 [23.89; 29.39] | 26.78 [24.34; 29.68] | 26.67 [24.23; 30.1] | 25.92 [24.19; 28.54] | 26.83 [25.07; 28.83] |
| Body Surface Area (m$^2$) Median[Q1;Q3] | 1.77 [1.65; 1.92] | 1.97 [1.81; 2.12] | 1.96 [1.82; 2.1] | 1.97 [1.82; 2.13] | 1.95 [1.79; 2.07] | 2.04 [1.87; 2.13] |
| **CMR** | Median[Q1;Q3] | Median[Q1;Q3] | Median[Q1;Q3] | Median[Q1;Q3] | Median[Q1;Q3] | Median[Q1;Q3] |
| Left ventricle end systolic volume (ml) | 31.07[26.62;36.41] | 33.6[27.75;41.24] | 34.77[28.48;43.15] | 38.55[29.92;50.25] | 35.47[28.54;42.46] | 49.67[40.72;62.56] |
| Left ventricle end diastolic volume (ml) | 78.57[70.4;87.33] | 79.76[69.56;90.04] | 81.85[71.76;93.8] | 83.47[71.16;97.44] | 79.54[72.3;92.18] | 96.14[79.36;112.76] |
| Left ventricle systolic volume (ml) | 47.13[41.91;52.97] | 45.31[38.38;52.15] | 46.21[40.27;51.93] | 44.21[38.5;50.6] | 45.86[41.3;52.71] | 44.44[39.25;51.54] |
| Left ventricle cardiac output (ml) | 5.05[4.36;5.86] | 5.37[4.57;6.31] | 5.2[4.51;6.07] | 5.31[4.53;6.28] | 5.44[4.61;6.12] | 5.45[4.64;6.39] |
| Left ventricle ejection fraction (%) | 60.08[56.44;63.76] | 57.55[51.63;62.33] | 57.06[51.48;61.68] | 53.57[46.25;60.24] | 57.53[51.43;63.45] | 45.75[39.05;51.42] |
| Left ventricle mass (g) | 42.54[38.03;48.4] | 47.52[41.29;54.41] | 49.7[43.84;55.29] | 50.66[43.88;58.27] | 56.93[48.13;71.86] | 53.77[47.17;62.73] |
| Right ventricle end systolic volume (ml) | 35.01[29.56;41.65] | 37.6[31.26;45.73] | 36.25[30.58;42.96] | 37.82[31.19;46.62] | 37.47[29.38;43.84] | 44.42[38.3;52.27] |
| Right ventricle end diastolic volume (ml) | 83.43[74.15;94.29] | 83.33[73.01;94.72] | 83.21[73.35;94.0] | 82.5[72.15;93.73] | 78.6[68.11;93.06] | 82.73[76.71;95.09] |
| Right ventricle systolic volume (ml) | 48.23[42.97;54.16] | 45.63[38.14;53.03] | 46.73[40.73;52.89] | 43.85[38.24;50.86] | 43.73[39.6;50.74] | 39.9[36.59;45.55] |
| Right ventricle ejection fraction (%) | 57.83[53.94;61.46] | 55.01[48.9;59.76] | 56.47[52.21;60.19] | 53.96[48.34;59.28] | 56.02[50.23;61.52] | 48.54[41.68;53.37] |
| Left atrium maximum volume (ml) | 37.93[31.54;44.96] | 46.01[35.94;58.52] | 40.41[32.55;49.8] | 44.86[34.54;56.73] | 44.86[37.99;62.56] | 50.38[38.55;58.53] |
| Left atrium minimum volume (ml) | 14.45[10.8;18.39] | 21.67[14.62;35.72] | 17.16[12.32;23.51] | 20.87[13.83;31.89] | 21.24[13.82;31.68] | 23.39[17.63;37.82] |
| Right atrium maximum volume (ml) | 46.98[39.79;55.64] | 51.29[40.88;65.43] | 44.18[36.44;54.34] | 46.22[38.14;61.14] | 47.83[36.1;62.0] | 52.11[42.01;61.79] |
| Right atrium minimum volume (ml) | 24.55[19.82;30.56] | 28.88[21.23;41.02] | 24.64[18.84;31.72] | 26.36[19.62;36.89] | 25.79[20.01;36.82] | 31.47[22.89;40.16] |
| Descending aorta distensibility ($10^{-3}$ .mmHg$^{-1}$) | 2.36[1.67;3.27] | 1.79[1.33;2.47] | 1.86[1.35;2.58] | 1.7[1.22;2.44] | 1.91[1.53;2.58] | 1.85[1.37;2.56] |
| Ascending aorta distensibility ($10^{-3}$ .mmHg$^{-1}$) | 1.69[1.12;2.56] | 1.29[0.91;1.96] | 1.32[0.94;1.94] | 1.28[0.88;1.9] | 1.33[0.97;1.72] | 1.35[0.99;2.0] |
| Global longitudinal strain (%) | -18.56[-20.25;-16.99] | -17.21[-19.51;-14.55] | -17.67[-19.5;-15.65] | -16.62[-18.49;-13.95] | -16.64[-19.18;-14.86] | -15.72[-17.34;-11.47] |
| Circumferential peak diastolic strain rate (s$^{-1}$) | 2.53[2.06;3.03] | 1.97[1.46;2.6] | 1.95[1.46;2.5] | 1.65[1.15;2.22] | 1.95[1.36;2.62] | 1.29[0.84;1.75] |
| Longitudinal peak diastolic strain rate (s$^{-1}$) | 1.75[1.4;2.14] | 1.48[1.11;1.9] | 1.46[1.14;1.86] | 1.29[0.98;1.68] | 1.33[1.13;1.95] | 1.12[0.89;1.48] |
| Radial peak diastolic strain rate (s$^{-1}$) | -6.23[-7.44;-5.07] | -5.17[-6.73;-3.81] | -5.09[-6.46;-3.75] | -4.31[-5.83;-2.99] | -5.0[-6.86;-3.49] | -3.02[-4.77;-2.09] |
| Maximum wall thickness (mm) | 8.59[7.67;9.57] | 10.25[9.13;11.41] | 10.22[9.27;11.34] | 10.39[9.22;11.6] | 12.45[11.17;15.21] | 10.43[9.15;12.05] |

**Supplementary Table 3. Positive and negative genes for HF.** The positive (n=143) and negative (n=143) genes were identified from the DISGENET database. HF: heart failure.

| Positive genes for HF | Negative genes for HF |
|---|---|
| ’PIK3IP1’, ’SERPINE1’, ’IL6’, ’FIBIN’, ’EDNRA’, ’KAT8’, ’ADRA1D’, ’PTH’, ’ALOX15’, ’SLC9A1’,’PTGS1’, ’IFNG’, ’CIDEA’, ’UCP1’, ’ATP2A1’, ’NPPA’, ’MYH6’, ’TNFRSF1A’, ’CYRIB’, ’WDR45’, ’APLN’, ’NRG1’, ’MS4A6A’, ’NOS2’, ’MSTN’, ’SIRT1’, ’PTPN3’, ’VEGFA’, ’ALB’, ’PTP4A2’, ’CXCL2’,’BAMBI’, ’INS’, ’APCS’, ’EPN3’, ’EDNRB’, ’ATP2A2’, ’RAC1’, ’HSPB1’, ’ACACA’, ’CFD’, ’POMC’, ’PTGS2’, ’ROCK2’, ’NR3C2’, ’GHRL’, ’SOD1’, ’PON1’, ’AGTR1’, ’PDPK1’, ’PIK3CG’, ’ADIPOQ’, ’PLAT’, ’EPHX2’, ’ADRB3’, ’HMOX1’, ’PCK1’, ’TNF’, ’TRDN’, ’FASN’, ’CXCL8’, ’NPPB’, ’AVP’, ’NUPR1’, ’SFRP1’, ’CSF2’, ’HAND2’, ’AGT’, ’PDGFRA’, ’TLR2’, ’APOC1’, ’FIP1L1’, ’FXYD3’,’ITGB1’, ’MYH7’, ’HAMP’, ’REN’, ’HTR2B’, ’PPP1R1A’, ’PEBP1’, ’CRP’, ’NPR1’, ’GCG’, ’SOD3’, ’CSF3’, ’CCN2’, ’COL4A1’, ’RETN’, ’PLXND1’, ’PPARG’, ’NRIP1’, ’FBLN5’, ’NOX4’, ’COL6A1’, ’SOX4’, ’CREG1’, ’ADRB1’, ’ACADS’, ’CAT’, ’ELOVL6’, ’AKIP1’, ’CYBB’, ’NFE2L2’, ’PRL’, ’ADRA2C’, ’NOS3’, ’XDH’,’CCL2’, ’ANKRD23’, ’EDN1’, ’COL8A1’, ’GRK2’, ’TNNT2’, ’ATP1A3’, ’GSK3B’, ’PPARGC1A’, ’ACLY’, ’AVPR2’, ’ACE’, ’KANK2’, ’CLIC2’, ’VWF’, ’PRKAR2B’, ’CDC25B’, ’FHL1’, ’TBX20’, ’CS’, ’GPX4’, ’SOD2’, ’RBP4’, ’IL1B’, ’OLR1’, ’DSTN’, ’SIPA1L1’, ’MAP2K7’, ’KLF9’, ’GDF15’, ’CXCL3’, ’UCN2’,’NOX1’, ’ELK3’, ’GATM’, ’HIF1A’ | ’HABP2’, ’CRACR2B’, ’RBMY2BP’, ’HINT1’, ’PSG2’, ’GPT’, ’ADAMDEC1’, ’IGKV1D-39’, ’NBAS’, ’PRDM12’,’AAGAB’, ’POC1B-GALNT4’, ’GID4’, ’OPA3’, ’TEP1’, ’UPP1’, ’GPHA2’, ’HINT3’, ’BTF3L4’, ’DNAL4’,’NSMCE1’, ’EP400P1’, ’LRRC8E’, ’FAM43A’, ’PEX13’, ’OR2H1’, ’PAGR1’, ’CORT’, ’NSA2’, ’ADAMTS4’, ’VWA8’, ’FGF23’, ’OPN1LW’, ’MGST1’, ’S1PR2’, ’TAS2R13’, ’WFDC9’, ’SPTLC3’, ’MAMDC2’, ’PAPOLG’, ’SWAP70’, ’NR2F1’, ’CHTF8’, ’KLHL10’, ’KCNMB2’, ’BCLAF3’, ’UNC79’, ’DLGAP3’, ’RNF115’, ’EMSY’,’SLC15A3’, ’CNPY1’, ’WASH6P’, ’CATSPERD’, ’ESCO2’, ’METTL27’, ’ITFG2’, ’SETD5’, ’ARMC12’, ’ST14’,’APCDD1L’, ’SCARNA27’, ’ANKRD53’, ’ZNF350’, ’SLC25A31’, ’GALNT6’, ’WFDC2’, ’IGHV4-31’, ’SMIM2’, ’ETHE1’, ’QRICH2’, ’PPFIBP2’, ’PCF11’, ’ALDH5A1’, ’SACS’, ’HRC’, ’C12orf29’, ’C9orf153’, ’SIKE1’,’COX6A2’, ’KLHL21’, ’FAM131A’, ’TAS2R7’, ’HOATZ’, ’CRYGS’, ’RFLNA’, ’GMEB1’, ’CYP46A1’, ’MIR425’, ’SMIM19’, ’TPTEP1’, ’METTL17’, ’MED12L’, ’SERF1A’, ’AGPAT5’, ’TMEM54’, ’PATE1’, ’KLHL3’, ’C3orf35’, ’ABTB1’, ’RPS6KC1’, ’ABCB7’, ’C16orf46’, ’SPATA13’, ’ZNF347’, ’STEAP4’, ’PCDH9’, ’FREM1’, ’AHSA1’,’SPATA1’, ’SOWAHC’, ’IGLV3-19’, ’IL17RD’, ’GRSF1’, ’CABCOCO1’, ’ABR’, ’ROBO1’, ’TLR7’, ’SH3BGR’,’CYP27C1’, ’KIF3B’, ’H2AZ2’, ’ESRRB’, ’LAD1’, ’PSORS1C3’, ’ASCL4’, ’TMIGD3’, ’OSBPL7’, ’CLP1’, ’HES5’, ’SELENOM’, ’IGLV3-12’, ’VAX2’, ’ARMC2’, ’FECH’, ’GSTT2’, ’TCN1’, ’PDZD11’, ’ALDOB’,’FAM72A’, ’GAGE5’, ’ENAH’, ’IGKV4-1’ |

**Supplementary Table 4. Positive and negative genes for AF.** The positive (n=160) and negative (n=160) genes were identified from the DISGENET database. AF: atrial fibrillation.

| Positive genes for AF | Negative genes for AF |
|---|---|
| ’GATA4’,’UBE4B’,’SCN10A’,’PPP2R3A’,’UST’,’AKAP6’, | SPATA13’, ’PRDM12’, ’VWA8’, ’PAGR1’, ’S1PR2’, ’CHAMP1’, |
| ’LHX3’,’CEP68’,’LINC00477’,’WNT3’,’PPFIA4’, | ’WFDC9’, ’EP400P1’, ’FAM131A’, ’KLHL3’, |
| ’PRRX1’,’WDR1’,’MBD5’,’IRF2BPL’,’SYNPO2L’, | ’AHSA1’, ’KCNMB2’, ’SLC25A31’, ’SIKE1’,’DUOXA1’, ’CHTF8’, |
| ’MYPN’,’TBX5’,’XPO1’,’HAND2’,’KCNN2’,’GOPC’, | ’GSTT2’, ’OSBPL7’, ’PCF11’, ’TLR7’,’AAGAB’, ’MGST1’, |
| ’TEX41’,’CASQ2’,’CAV2’,’NPPB’,’RPL3L’,’KCNN3’, | ’MAMDC2’, ’DNAL4’, ’VAX2’, ’IGKV4-1’, ’CLP1’, ’UNC79’, ’CYP46A1’, |
| ’SSPN’,’SPATS2L’,’PITX2’,’NEURL1’,’GCOM1’,’VWF’, | ’CRYGS’,’CRACR2B’, ’ZNF347’, ’NR2F1’, ’ADAMTS4’, ’ISCU’, |
| ’LRIG1’,’KDM1B’,’PHLDB2’,’PLN’,’TNFSF12’,’SLC9B1’, | ’FAM72A’, ’AGPAT5’, ’KIF3B’, ’CABCOCO1’, ’ARMC2’,’SMIM2’, |
| ’NKX25’,’KIF3C’,’KCNJ8’,’RPS2’,’CAV1’,’ARHGAP10’, | ’CALCOCO1’, ’ETHE1’,’HRC’, ’PEX13’, ’SH3BGR’, ’NFASC’, |
| ’HIP1R’,’HCN4’,’PKP2’,’IGF1R’,’ABCC9’,’SOX5’, | ’CYP27C1’, ’MED12L’, ’NBAS’,’SOWAHC’, ’ALDOB’, ’ABTB1’, |
| ’DNAH10’,’KCNE5’,’LRMDA’,’SMAD7’,’SCMH1’,’TTN’, | ’TMIGD3’, ’FREM1’, ’BBIP1’, ’PATE1’, ’GMEB1’, |
| ’HSF2’,’MYH6’,’ATXN1’,’CREB5’,’MYOCD’,’PSMB7’, | ’OPA3’, ’TAS2R13’, ’ABR’,’C5orf34’, ’ARMC12’, ’RFLNA’, |
| ’ARNT2’,’ASAH1’,’TUBA8’,’NR3C1’,’AGBL4’,’AOPEP’, | ’TCN1’, ’PDZD11’, ’SWAP70’, ’TEP1’, ’COX6A2’, ’HINT1’, |
| ’NACA’,’DGKB’,’MYOT’,’SGCA’,’CYTH1’, | ’SCARNA27’,’HINT3’, ’TTC8’, ’PSG2’, ’DHRS9’, ’ITFG2’, |
| ’REEP1’,’CAND2’,’MYH7’,’SYNE2’,’GTF2I’,’SH3PXD2A’, | ’HABP2’, ’USB1’, ’NSMCE1’, ’TAS2R7’, ’SLITRK4’, ’CNPY1’, |
| ’COG5’,’RBM20’,’MYO1C’,’HERC1’,’LINC01426’,’CASZ1’, | ’C9orf153’, ’IGHV4-31’, ’POC1B-GALNT4’, ’METTL17’, ’TUBD1’, ’ESCO2’, |
| ’FBRSL1’,’WNT8A’,’MYOZ1’,’WIPF1’,’CUL4A’,’CGA’, | ’TMEM54’, ’ANKRD53’,’FECH’,’IGLV3-12’, ’CORT’, ’GPHA2’, |
| ’SLC35F1’,’CDKN1A’,’SCN5A’,’USP3’,’C1orf185’,’XPO7’, | ’C12orf29’, ’MED11’, ’BTF3L4’,’STEAP4’, ’MIR425’, ’C16orf46’, |
| ’PHLDA1’,’KCNJ5’,’ZPBP2’,’GNB4’,’OPN1SW’,’XXYLT1’, | ’WNT3A’,’C3orf35’, ’TPTEP1’, ’LAD1’, ’SETD5’, ’OR2H1’, |
| ’USP34’,’ZFHX3’,’MIR11HG’,’ARMH3’,’FGF5’,’FRMD4B’, | ’PAXX’, ’GAGE5’, ’GID4’, ’ZNF350’, ’PSORS1C3’, |
| ’SORL1’,’CYP2C19’,’NOS3’,’CDK6’,’HSPG2’,’LRRC10’, | ’IGKV1D-39’, ’RPS6KC1’, ’NSA2’, ’RBMY2BP’,’ROBO1’, ’METTL27’, |
| ’LINC01142’,’HTR4’,’ERBB4’,’SLIT3’,’THRB’,’MIR6500’, | ’SMIM19’, ’ASCL4’, ’HOATZ’,’APCDD1L’, ’KLHL21’, ’SPATA1’ |
| ’KCNA5’,’GOSR2’,’SLC27A6’,’SNRNP27’,’KCNH2’,’KCNE2’, | , ’FAM43A’,’GALNT6’, ’GRSF1’, ’ST14’, ’DLGAP3’, ’FGF23’, |
| ’NUCKS1’,’KCNJ2’,’PTK2’,’PAK2’, | ’GPT’, ’QRICH2’,’SACS’, ’ADAMDEC1’, ’ALDH5A1’, ’IGLV3-19’, |
| ’SIRT1’,’DPF3’,’SOX15’,’ZNF462’,’SUN1’,’CFL2’, | ’UPP1’, ’ABCB7’, ’GLP2R’, ’SELENOM’, ’ESRRB’, ’WASH6P’, |
| ’SELE’,’CAMK2D’,’ACE’,’DES’,’REEP3’,’BEST3’, | ’OPN1LW’, ’KLHL10’, ’IL17RD’, ’HES5’,’WFDC2’,’PCDH9’, |
| ’CALU’,’FBXO32’,’EPHA3’,’MYO18B’,’NEBL’,’MAPT’, | ’AK8’,’EMSY’, ’ENAH’, ’SERF1A’, ’H2AZ2’, ’PPFIBP2’, |
| ’KCND3’,’GYPC’,’ESR2’,’EDN1’,’TTNAS1’,’MIR30B’, | ’RNF115’,’SLC15A3’,’CATSPERD’, ’SPTLC3’, ’LRRC8E’, ’PAPOLG’, |
| ’NAV2’,’MEX3C’,’GJA5’ | ’BCLAF3’ |

**Supplementary Table 5. Positive and negative genes for MI.** The positive (n=95) and negative (n=90) genes were identified from the DISGENET database. MI: myocardial infarction.

| Positive genes for MI | Negative genes for MI |
|---|---|
| ’LDHA’, ’MIR145’, ’NR3C2’, ’NPPA’, ’TNNI3’,’AGT’, | ’SPATA13’, ’PRDM12’, ’VWA8’, ’PAGR1’, ’S1PR2’, ’CHAMP1’, |
| ’PPARGC1A’, ’EPO’, ’MYH7’, ’BRAP’, ’PDE3A’, ’SH2B3’, | ’WFDC9’, ’EP400P1’, ’FAM131A’, ’KLHL3’,’AHSA1’, ’KCNMB2’, |
| ’S100B’, ’BAK1’, ’SOD2’, ’KLF13’, ’BECN1’, ’ADORA1’, | ’SLC25A31’, ’SIKE1’, ’DUOXA1’, ’CHTF8’, ’GSTT2’, ’OSBPL7’, |
| ’F13A1’, ’DGKZ’, ’MIR125B2’, ’BCL2’,’APOE’, ’TGFB1’, | ’PCF11’, ’TLR7’,’AAGAB’, ’MGST1’, ’MAMDC2’, ’DNAL4’, ’VAX2’, |
| ’NOS3’, ’P2RY12’, ’ATM’, ’MFF’, ’GGT1’, ’LRP8’, ’MMP2’, | ’IGKV4-1’, ’CLP1’, ’UNC79’, ’CYP46A1’, ’CRYGS’,’CRACR2B’, |
| ’TM6SF2’,’PHACTR1’, ’F2’, ’GSK3B’, ’WDR12’, ’NOS2’, | ’ZNF347’, ’NR2F1’, ’ADAMTS4’, ’ISCU’, ’FAM72A’, ’AGPAT5’, |
| ’ACTA2’, ’BCL2L1’, ’ICAM1’, ’PSMA6’, ’MMP9’,’ADORA3’, | ’KIF3B’, ’CABCOCO1’, ’ARMC2’, ’SMIM2’, ’CALCOCO1’, |
| ’DDIT3’,’REN’, ’SOD1’, ’PLAT’, ’LGALS3’, ’IL1RN’, ’IL1B’, | ’ETHE1’, ’HRC’, ’PEX13’, ’SH3BGR’, ’NFASC’, ’CYP27C1’, |
| ’ITGB3’, ’HSPA5’,’PLAU’, ’SELL’, ’CASP3’, ’F5’, ’PAPPA’, | ’MED12L’, ’NBAS’,’SOWAHC’, ’ALDOB’, ’ABTB1’, ’TMIGD3’, |
| ’NRIP1’, ’ESR1’, ’THBD’,’LTA’, ’CKM’, ’HP’,’F7’, ’ATP1A1’, | ’FREM1’, ’BBIP1’, ’PATE1’, ’GMEB1’, ’OPA3’, ’TAS2R13’, ’ABR’, |
| ’CKB’, ’CAT’, ’MIA3’,’PRKCE’, ’GATA4’, ’GCLM’,’KLK1’, | ’C5orf34’, ’ARMC12’, ’RFLNA’, ’TCN1’, ’PDZD11’, ’SWAP70’, |
| ’HMGB1’, ’KIF6’, ’HSD11B2’,’MIR423’, ’TNF’, ’IL10’, | ’TEP1’, ’COX6A2’, ’HINT1’, ’SCARNA27’,’HINT3’, ’TTC8’, |
| ’OLR1’, ’LGALS2’, ’MIR761’, ’IFNA2’,’BAX’, ’ACE’, | ’PSG2’, ’DHRS9’, ’ITFG2’, ’HABP2’, ’USB1’, ’NSMCE1’, ’TAS2R7’, |
| ’MIAT’, ’GCLC’, ’CREB1’, ’CREM’,’PLN’, ’TNFSF4’ | ’SLITRK4’, ’CNPY1’,’C9orf153’, ’IGHV4-31’, ’POC1B-GALNT4’, |
| | ’METTL17’, ’TUBD1’, ’ESCO2’, ’TMEM54’, ’ANKRD53’ |

**Supplementary Table 6. The performance of the ML classifiers in predicting gene-disease associations.** The accuracy (Acc) and area under the curve (AUC) for predicting the links between the genes and the diseases using three machine learning classifiers including random forest (RF), support vector machine (SVM) and artificial neural networks (ANNs). CMR:cardiac magnetic resonance imaging, AF: atrial fibrillation, MI: myocardial infarction, HF: heart failure.

| Disease | Classifier | CMR | | NO CMR | |
|---|---|---|---|---|---|
| | | Acc | AUC | Acc | AUC |
| HF | SVM | 72.4% | 0.80 | 70.6% | 0.79 |
| | RF | 62.1% | 0.72 | 65.5% | 0.70 |
| | ANN | 69.0% | 0.80 | 70.6% | 0.79 |
| AF | SVM | 75.0% | 0.78 | 75.0% | 0.76 |
| | RF | 71.9% | 0.75 | 70.3% | 0.75 |
| | ANN | 73.4% | 0.78 | 68.3% | 0.78 |
| MI | SVM | 83.3% | 0.83 | 81.1% | 0.82 |
| | RF | 75.6% | 0.79 | 81.0% | 0.82 |
| | ANN | 83.7% | 0.82 | 70.2% | 0.83 |

**Supplementary Table 7. Top 10 predicted genes and their probabilities for each disease with and without CMR features.** CMR:cardiac magnetic resonance imaging, HF: heart failure, AF: atrial fibrillation, MI: myocardial infarction.

| HF | | | | AF | | | | MI | | | |
|---|---|---|---|---|---|---|---|---|---|---|---|
| CMR | | NO-CMR | | CMR | | NO-CMR | | CMR | | NO-CMR | |
| Gene | Probability | Gene | Probability | Gene | Probability | Gene | Probability | Gene | Probability | Gene | Probability |
| EGR1 | 0.99 | PRKCZ | 0.98 | DAPK1 | 0.99 | CLTC | 0.99 | SUMO1 | 0.99 | MAFF | 0.99 |
| CTCF | 0.99 | ZNF417 | 0.98 | SRC | 0.99 | SNW1 | 0.99 | TCF12 | 0.99 | ZBTB7A | 0.99 |
| ETS1 | 0.99 | NEDD8 | 0.98 | EP300 | 0.99 | CBL | 0.99 | H2AX | 0.99 | SGTB | 0.99 |
| GATA2 | 0.99 | MTA1 | 0.98 | COPS5 | 0.99 | HNRNPA1 | 0.99 | RAD21 | 0.99 | FOXA2 | 0.99 |
| APP | 0.99 | ZBTB16 | 0.98 | GATA1 | 0.99 | ILF3 | 0.99 | GATA1 | 0.99 | SREBF1 | 0.99 |
| MYC | 0.99 | CDK9 | 0.98 | H2AX | 0.99 | PDE4DIP | 0.99 | USF2 | 0.99 | BATF | 0.99 |
| EP300 | 0.99 | PRPF31 | 0.98 | HSPA8 | 0.99 | HNRNPK | 0.99 | HTT | 0.99 | EFNA5 | 0.99 |
| RAD21 | 0.99 | HNRNPM | 0.98 | PDHA1 | 0.99 | EED | 0.99 | PCNA | 0.99 | KRTAP10-3 | 0.99 |
| AR | 0.99 | DNM1L | 0.98 | CASP8 | 0.99 | APC | 0.99 | UBL4A | 0.99 | SP2 | 0.99 |
| UBC | 0.99 | SNRNP70 | 0.98 | HNF4A | 0.99 | NFKBIA | 0.99 | SNCA | 0.99 | APPBP2 | 0.99 |

**Supplementary Table 8. Identified candidate medications and their target for repurposing in heart failure.**

| Rank | Medication | ATC code | Target genes | Predicted score | Supporting evidence | Known indications |
|---|---|---|---|---|---|---|
| 1 | Glutamic acid | A09AB01 | NAGS, GRIN3A, CPQ, EARS2, SLC1A2, GRIN2C, SLC1A7, GRM4, FOLH1, DNPEP, GRIN2D, GLUD2, AASS, GRIK2, GRIK4, AADAT, GRM7, ENPEP, SLC1A6, BCAT2, GRIK1, GOT2, SLC7A11, GRM8, SLC1A3, GCLC, GPT, GMPS, GPT2, GOT1, GRIA4, NADSYN1, GRIA3, PSAT1, FPGS, GRIN2A, ABAT, GRIK3, BCAT1, GRIA2, GRIA1, GRIN1, GRIK5, TAT, GRM1, EPRS1, ALDH18A1, GLS, GAD2, SLC1A1, GGCX, GLS2, GAD1, ASNS, FTCD, GCLM, OPLAH, GRIN2B, GLUL, GLUD1, PFAS | 0.98 | response to oxidative stress, regulation of neuron apoptotic process | helps speed the healing of ulcers and helps control schizophrenia |
| 2 | Methotrexate | L04AX03 | ATIC, TYMS, DHFR | 0.97 | regulation of response to oxidative stress, regulation of cellular response to oxidative stress | treats cancer, autoimmune diseases and rheumatoid arthritis |
| 3 | Topiramate | N03AX11 | SCN7A, SCN10A, SCN9A, SCN11A, SCN8A, SCN1A, SCN3A, CACNA1F, SCN4A, CACNB1, GRIK2, GRIK4, CACNA1D, CA3, GRIK1, CA4, SCN2A, CA1, CACNB2, CACNB3, CACNA1E, CACNA1C, CA2, GRIK3, CACNB4, CACNA1S, SCN5A, GRIK5, GABRA1 | 0.96 | regulation of heart rate by cardiac conduction, regulation of ventricular cardiac muscle cell action potential | antiepileptic |
| 4 | Probenecid | M04AB01 | TAS2R16, SLC22A11, SLC22A8, PANX1, SLC22A6 | 0.96 | positive regulation of macrophage cytokine production | treats chronic gout or gouty arthritis |
| 5 | Estradiol acetate | G03CA03 | HSD17B2, MT-ATP6, GPER1, NR1I2, BECN1, NCOA2, CHRNA4, ESR2, ESR1, ESRRG | 0.96 | positive regulation of cardiac vascular smooth muscle cell differentiation, positive regulation of apoptotic signaling pathway | reduces symptoms of menopause |
| 6 | Caffeine | N06BC01 | PDE7B, PDE8B, PDE11A, PDE6B, PDE1C, PDE1A, PDE5A, PDE4B, ADORA3, PDE6A, PDE10A, ADORA2B, PDE6C, PDE4A, ADORA2A, PDE4C, PDE1B, PDE2A, PIK3CD, PDE8A, ADORA1, PIK3CB, PDE9A, PRKDC, ITPR3, ITPR2, PDE3B, PIK3CA, PDE7A, PDE3A, RYR1, PDE4D, ITPR1, ATM | 0.95 | regulation of cardiac muscle cell contraction, heart contraction | pain relief product |
| 7 | Cannabidiol | N03AX24 | TRPV2, TRPV3, TRPV4, TRPA1, GPR55, PTGS1, CACNA1G, HTR1A, GLRA3, CACNA1H, OPRD1, CYP1B1, TRPV1, CYP1A2, CACNA1I, NQO1, CYP3A5, GPR18, NAAA, CYP2D6, CYP17A1, CYP3A7, GPX1, GPR12, CNR2, CAT, IDO1, ACAT1, ADORA1, TRPM8, CNR1, CHRNA7, HMGCR, GLRA1, GSR, OPRM1, HTR2A, VDAC1, PPARG, PTGS2, SOD1, GLRB, HTR3A | 0.95 | regulation of heart contraction, cellular response to oxidative stress | treats certain types of epilepsy such as Lennox-Gastaut syndrome |
| 8 | Prasterone | A14AA07 | GABRP, GABRG3, GRIN3A, GRIN3B, GABRQ, GRIN2C, SIGMAR1, GABRA5, GABRA2, GABRB1, GRIN2D, GABRA4, GABRG1, GRIN2A, GABRG2, GABRD, GABRB2, GABRE, GRIN1, GABRA3, NR1I2, NR1I3, PPARA, GABRB3, AR, GABRA6, GRIN2B, ESR2, GABRA1, ESR1 | 0.95 | regulation of epithelial cell apoptotic process | improves symptoms of lupus and erection in men |
| 9 | Glycine | B05CX03 | GLYATL2, GLYATL1, AGXT2, GRIN3B, GRIN2C, GLRA3, GLYAT, GATM, GPR18, GRIN2A, PIPOX, GLRA2, ALAS2, SHMT1, GCAT, GNMT, GARS1, GLRA1, ALAS1, GSS, BAAT, SHMT2, GLRB, AGXT | 0.95 | mitochondrial gene expression | antioxidant and anti-inflammatory |
| 10 | Ranolazine | C01EB18 | KCNJ14, SCN7A, SCN10A, SCN9A, SCN11A, SCN8A, SCN1A, SCN3A, SCN4B, SCN1B, CACNA1F, SCN4A, KCNJ12, CACNB1, KCNJ4, CACNA1D, ADRA1D, SCN2A, CACNB2, CACNB3, CACNA1C, ADRA1B, ADRA1A, CACNB4, CACNA1S, SCN5A, SCN2B, KCNJ2, SCN3B, ADRB1 | 0.95 | regulation of ventricular cardiac muscle cell action potential, positive regulation of heart rate by epinephrine-norepinephrine | treats chest pain related to heart |

**Supplementary Table 9. The results of validating the methotrexate in heart failure using the survival model.** n is the number of patients taking the medciation. The baseline is heart failure with rheumatoid arthritis.

| Medication | n | Sex (female%) | Ethnicity (white%) | Age Median[Q1;Q3] | BMI Median[Q1;Q3] |
|---|---|---|---|---|---|
| Methotrexate | 147 | 81 (55.1%) | 141 (95.9%) | 68[62;74] | 28.4[24.5;32.2] |
| Sulfasalazine | 40 | 19 (47.5%) | 40 (100%) | 68.5[61;72.25] | 28.6[24.8;32.8] |
| Hydroxychloroquine | 47 | 33 (70.2%) | 45 (95.7%) | 67[61.5;73] | 29.4[25.2;35.6] |

**Supplementary Table 10. The survival probability and hazard ratio for the predicted medication for heart failure (HF).** The calculate survival probability at 5 years and at 10 years. The extracted cohort includes individuals with HF and rheumatoid arthritis.

| Group | HR [95% CI] | Survival probbility at 5 years [95% CI] | Survival probbility at 10 years [95% CI] | p value |
|---|---|---|---|---|
| Methotrexate (Baseline, n=147) | 1.0 | 0.75 [0.67, 0.84] | 0.62 [0.50, 0.74] | NA |
| Hydroxychloroquine (n=47) | 1.76 [1.11, 2.79] | 0.64 [0.51, 0.79] | 0.48 [0.31, 0.67] | 0.02 |
| Sulfasalazine (n=40) | 1.63 [0.98, 2.72] | 0.67 [0.57, 0.77] | 0.52 [0.38, 0.64] | 0.06 |

**Supplementary Table 11. Identified candidate medications and their target for repurposing in atrial fibrillation.**

| Rank | Medication | ATC code | Target genes | Predicted score | Supporting evidence | Known indications |
|---|---|---|---|---|---|---|
| 1 | Methotrexate | L04AX03 | ATIC, TYMS, DHFR | 0.80 | regulation of response to oxidativestress, regulation of cellular response to oxidative stress | Treats cancer, autoimmune diseases and rheumatoid arthritis |
| 2 | Zonisamide | N03AX15 | CA7, CA5B, CA13, SCN9A, SCN11A, CACNA1G, SCN1A, SCN3A, SCN4B, SCN1B, CACNA1H, SCN4A, CA11, CACNA1I, CA3, CA5A, CA12, CA4, SCN2A, CA1, CA14, CA6, CA10, CA2, CA9, MAOA, MAOB, SCN5A, SCN2B, CA8, SCN3B | 0.80 | regulation of ventricular cardiac muscle cell membrane repolarization, regulation of ventricular cardiac muscle cell membrane depolarization | antiepileptic |
| 3 | Probenecid | M04AB01 | TAS2R16, SLC22A11, SLC22A8, PANX1, SLC22A6 | 0.79 | positive regulation of macrophage cytokine production | treats chronic gout or gouty arthritis |
| 4 | Estradiol valerate | G03CA03 | HSD17B2, MT-ATP6, GPER1, NR1I2, BECN1, NCOA2, CHRNA4, ESR2, ESR1, ESRRG | 0.79 | positive regulation of cardiac vascular smooth muscle cell differentiation, positive regulation of apoptotic signaling pathway | treating conditions related to low estrogen levels, such as menopausal symptoms and certain cancers |
| 5 | Estradiol cypionate | G03CA03 | HSD17B2, MT-ATP6, GPER1, NR1I2, BECN1, NCOA2, CHRNA4, ESR2, ESR1, ESRRG | 0.79 | positive regulation of cardiac vascular smooth muscle cell differentiation, positive regulation of apoptotic signaling pathway | treatment of hypoestrogenism |
| 6 | Estradiol acetate | G03CA03 | HSD17B2, MT-ATP6, GPER1, NR1I2, BECN1, NCOA2, CHRNA4, ESR2, ESR1, ESRRG | 0.79 | cardiac vascular smooth muscle cell differentiation, positive regulation of apoptotic signaling pathway | reduces symptoms of menopause |
| 7 | Glycine | B05CX03 | GLYATL2, GLYATL1, AGXT2, GRIN3B, GRIN2C,GLRA3, GLYAT, GATM, GPR18, GRIN2A, PIPOX, GLRA2, ALAS2, SHMT1, GCAT, GNMT, GARS1,GLRA1, ALAS1, GSS, BAAT, SHMT2, GLRB, AGXT | 0.77 | mitochondrial gene expression | used as irrigation during surgery |
| 8 | Glutamic acid | A09AB01 | NAGS, GRIN3A, CPQ, EARS2, SLC1A2, GRIN2C,SLC1A7, GRM4, FOLH1, DNPEP, GRIN2D, GLUD2,AASS, GRIK2, GRIK4, AADAT, GRM7, ENPEP,SLC1A6, BCAT2, GRIK1, GOT2, SLC7A11, GRM8, SLC1A3, GCLC, GPT, GMPS, GPT2, GOT1, GRIA4, NADSYN1, GRIA3, PSAT1, FPGS, GRIN2A, ABAT, GRIK3, BCAT1, GRIA2, GRIA1, GRIN1, GRIK5, TAT,GRM1, EPRS1, ALDH18A1, GLS, GAD2, SLC1A1, GGCX, GLS2, GAD1, ASNS, FTCD, GCLM, OPLAH,GRIN2B, GLUL, GLUD1, PFAS | 0.76 | response to oxidative stress, regulation of apoptotic process | helps speed the healing of ulcers and helps control schizophrenia |
| 9 | Acamprosate | N07BB04 | GABRP, GABRG3, GRIN3A, GRIN3B, GABRQ, GRIN2C, GABRA5, GABRA2, GABRB1, GRIN2D, GABRA4, GABRG1, GRIN2A, GABRG2, GABRD, GRM5, GABRB2, GABRE, GRIN1, GABRA3 | 0.76 | abnormal nervous system electrophysiology, positive regulation of cell communication | reduces alcoholism cravings |
| 10 | Pentobarbital | N05CA01 | GABRP, GABRG3, GRIN3A, GRIN3B, GABRQ, GRIN2C, GABRA5, GABRA2, GABRB1, GRIN2D, GRIK2, GABRA4, GABRG1, GRIN2A, GABRG2, GABRD, GRIA2, GABRB2, GABRE, GRIN1, GABRA3, CHRNA7, NR1I2, GABRB3, CHRNA4, GABRA6, GRIN2B, GABRA1 | 0.75 | positive regulation of cell communication, abnormal nervous system electrophysiology | treate seizures |

**Supplementary Table 12. The results of validating the methotrexate in atrial fibrillation using the survival model.** n is the number of patients taking the medciation. The baseline is atrial fibrillation with rheumatoid arthritis.

| Medication | n | Sex (female%) | Ethnicity (white%) | Age Median[Q1;Q3] | BMI Median[Q1;Q3] |
|---|---|---|---|---|---|
| Methotrexate | 170 | 79 (57.1%) | 167 (98.2%) | 71.0[64.0;75.6] | 28.0[25.4;31.9] |
| Sulfasalazine | 46 | 23 (50.0%) | 45 (97.8%) | 71.0[67.0;76.0] | 28.7[24.6;31.9] |
| Hydroxychloroquine | 39 | 29 (74.4%) | 37 (94.9%) | 69.0[66.5;75.0] | 30.9[26.1;39.2] |

**Supplementary Table 13. The survival probability and hazard ratio for the predicted medication for atrial fibrillation (AF).** The calculate survival probability at 5 years and at 10 years. The extracted cohort includes individuals with AF and rheumatoid arthritis.

| Group | HR [95% CI] | Survival probbility at 5 years [95% CI] | Survival probbility at 10 years [95% CI] | p value |
|---|---|---|---|---|
| Methotrexate (Baseline, n= 170) | 1.0 | 0.85 [0.79, 0.94] | 0.70 [0.58, 0.86] | NA |
| Hydroxychloroquine (n=39) | 1.76 [0.89, 3.49] | 0.75 [0.65, 0.86] | 0.54 [0.35, 0.69] | 0.10 |
| Sulfasalazine (n=46) | 1.01 [0.48, 2.10] | 0.84 [0.78, 0.92] | 0.68 [0.55, 0.82] | 0.90 |

**Supplementary Table 14. Identified candidate medications and their target for repurposing in mayocardial infarction.**

| Rank | Medication | ATC code | Target genes | Predicted score | Supporting evidence | Known indications |
|---|---|---|---|---|---|---|
| 1 | Vorinostat | L01XH01 | HDAC8, HDAC2, HDAC6,HDAC1, HDAC3 | 0.99 | positive regulation of response to oxidative stress, negative regulation of programmed cell death | treats cutaneous T-cell lymphoma (CTCL) |
| 2 | Podofilox | D06BB04 | TUBB,TUBA4A, TOP2A | 0.99 | cellular component disassembly involved in execution phase of apoptosis | treats molluscum contagiosum |
| 3 | Mecasermin rinfabate | H01AC05 | IGF2R, IGF1R, INSR | 0.99 | regulation of programmed cell death, heart development | treats growth failure in children with severe primary IGF-1 deficiency |
| 4 | Ingenol mebutate | D06BX02 | PRKCD,PRKCA | 0.99 | intrinsic apoptotic signaling pathway in response to oxidative stress, negative regulation of inflammatory response | treats actinic keratosis |
| 5 | Dimethyl fumarate | L04AX07 | RELA,KEAP1 | 0.99 | negative regulation of extrinsic apoptotic signaling pathway, cellular response to oxidative stress | treats the relapsing forms of multiple sclerosis |
| 6 | Glucosamine | M01AX05 | MMP9,IFNG NFKB2,TNF | 0.99 | extrinsic apoptotic signaling pathway via death domain receptors, apoptotic mitochondrial changes | might provide some pain relief for people with osteoarthritis |
| 7 | Lasofoxifene | G03XC03 | CNR2,ESR2, ESR1 | 0.99 | inflammatory response, negative regulation of muscle cell apoptotic process | prevention and treatment of osteoporosis |
| 8 | Dexrazoxane | V03AF02 | TOP2B, TOP2A | 0.99 | execution phase of apoptosis, cellular component disassembly involved in execution phase of apoptosis | decreases damage to the skin and tissues that may be caused when an anthracycline chemotherapy medication |
| 9 | Omega-3 | C10AX06 | PPARA,PPARG, SREBF1 | 0.99 | regulation of apoptotic process, adaptation cardiac muscle tissue growth, regulation of cardiac muscle tissue growth, cardiac muscle cell development | decreases the amount of triglycerides and other fats made in the liver |
| 10 | Auranofin | M01CB03 | PRDX5, IKBKB | 0.99 | inflammatory response | treats rheumatoid arthritis |

**Supplementary Table 15. Medications used to train the machine learning model for predicting new associations between medications and heart failure.**

| Medication indicated for the treatment of HF | Medication contraindicated for the treatment of HF |
|---|---|
| "Adomiparin", "Parnaparin", "Evolocumab", "Valsartan", "Bivalirudin", "Prasugrel", "Esmolol", "Fondaparinux", "Reteplase", "Nitroglycerin", "Phenprocoumon", "Alteplase", "Ardeparin", "Irbesartan", "Eptifibatide", "Tirofiban", "Dalteparin", "Dicoumarol", "Abciximab","Tenecteplase", "Heparin", "Alirocumab", "Atenolol", "Morphine", "Ticagrelor", "Vorapaxar", "Candesartan", "Captopril", "Lisinopril", "Quinapril", "Acetylsalicylic acid", "Enalapril", "Metoprolol", "Clopidogrel", "Losartan", "Heparin" | "Metamfetamine", "Testosterone cypionate", "Testosterone enanthate", "Sildenafil", "Saxagliptin", "Levodopa", "Furosemide", "Phenobarbital", "Guanfacine", "Flecainide", "Indomethacin", "Lovastatin", "Progesterone", "Testosterone", "Simvastatin", "Dipyridamole", "Pioglitazone", "Nefazodone", "Paclitaxel", "Phenytoin", "Liothyronine", "Levothyroxine", "Diclofenac", "Estradiol", "Meclofenamic acid", "Estradiol cypionate", "Estradiol valerate", "Digoxin", "Ribavirin", "Inositol nicotinate", "Levomefolic acid", "Acyclovir", "Sorafenib", "Metformin", "Sodium citrate", "Inositol" |

**Supplementary Table 16. Medications used to train the machine learning model for predicting new associations between medications and atrila fibrillation.**

| Medication indicated for the treatment of AF | Medication contraindicated for the treatment of AF |
|---|---|
| "Dabigatran", "Sotalol", "Warfarin", "Heparin", "Atenolol", "Diltiazem", "Timolol", "Bisoprolol", "Carvedilol", "Propafenone", "Edoxaban", "Betrixaban", "Rivaroxaban", "Apixaban", "Labetalol", "Dofetilide", "Metoprolol", "Propranolol", "Amiodarone", "Clopidogrel", "Quinidine", "Flecainide", "Dronedarone", "Digoxin", "Verapamil", "Procainamide", "Nadolol","Dipyridamole","Esmolol","Acetyldigitoxin" | "Bupivacaine", "Methylphenidate", "Cholecalciferol", "Lacosamide", "Alendronic acid", "Dobutamine","Ergocalciferol", "Lidocaine", " Fidaxomicin", "Azithromycin", "Fusidic acid", "Amoxicillin", "Naproxen", "Ibuprofen", "Oxytetracycline", "Tetracycline", "Disopyramide", "Quinine", "Epinephrine", "Conivaptan","Cetirizine", "Dorzolamide","Pantoprazole" ,"Loratadine","Raltitrexed","Methadone", "Erythromycin", "Clarithromycin" |

**Supplementary Table 17. Medications used to train the machine learning model for predicting new associations between medications and myocardial infarction.**

| Medication indicated for the treatment of MI | Medication contraindicated for the treatment of MI |
|---|---|
| "Adomiparin", "Heparin", "Parnaparin", "Evolocumab", "Bivalirudin", "Prasugrel", "Esmolol", "Fondaparinux", "Reteplase","Nitroglycerin", "Phenprocoumon", "Alteplase", "Ardeparin", "Irbesartan", "Eptifibatide", "Tirofiban", "Dalteparin", "Dicoumarol", "Abciximab","Tenecteplase", "Heparin", "Alirocumab", "Atenolol", "Morphine", "Ticagrelor", "Vorapaxar", "Candesartan", "Captopril", "Lisinopril", "Quinapril", "Acetylsalicylic acid", "Enalapril", "Metoprolol", "Clopidogrel", "Valsartan", "Losartan" | "Metamfetamine", "Testosterone cypionate", "Testosterone enanthate", "Sildenafil", "Saxagliptin", "Levodopa", "Furosemide", "Phenobarbital", "Guanfacine", "Flecainide", "Indomethacin", "Lovastatin", "Progesterone", "Testosterone", "Simvastatin", "Dipyridamole", "Pioglitazone", "Nefazodone", "Paclitaxel", "Phenytoin", "Liothyronine", "Levothyroxine", "Diclofenac", "Estradiol", "Meclofenamic acid", "Estradiol cypionate", "Estradiol valerate", "Digoxin", "Ribavirin", "Inositol nicotinate", "Levomefolic acid", "Acyclovir", "Sorafenib", "Metformin", "Sodium citrate", "Inositol" |

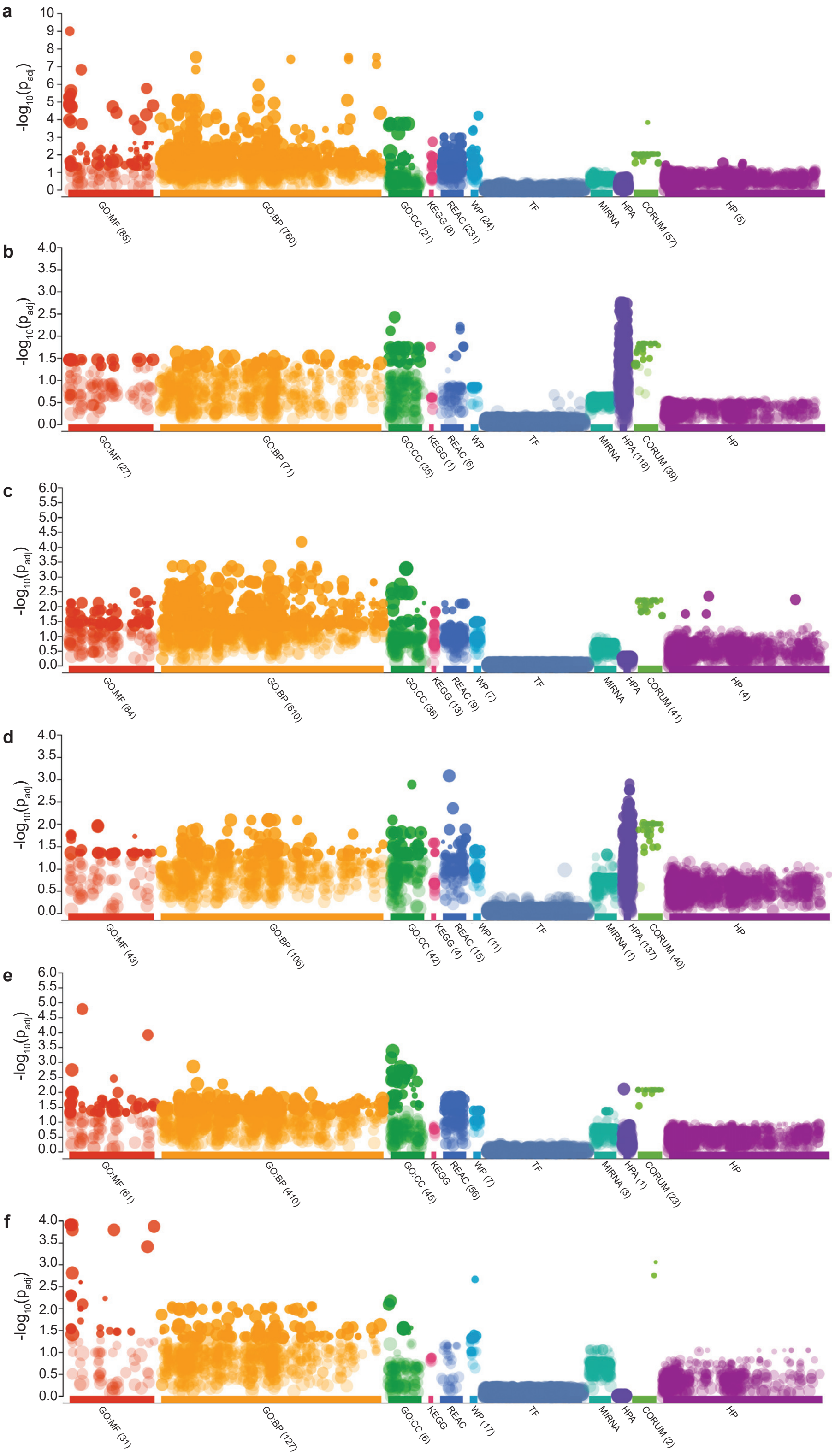


**Supplementary Figure 1. The enrichment analysis for the top ten predicted genes in the diseases of interest**. gprofiler has been used to perform the enrichment analysis and to generate the figures. x-axis represents the selected databases for the enrichment analysis and y-axis represents the adjusted p values using Benjamini-Hochberg (BH). **a:** is the enrichment analysis for heart failure (HF) using the predicted genes with integrating the cardiac magnetic resonance (CMR) data. **b:** is the enrichment analysis for HF using the predicted genes without the CMR. **c:** is the enrichment analysis for atrial fibrillation (AF) using the predicted genes with integrating the CMR. **d:** Shows the enrichment analysis for AF using the predicted genes without the CMR. **e:** is the enrichment analysis for myocardial infarction (MI) using the predicted genes with integrating the CMR. **f:** Shows the enrichment analysis for MI using the predicted genes without the CMR. GO: gene ontology, MF: molecular function, BP: biological process, CC: cellular component, KEGG: Kyoto Encyclopedia of Genes and Genomes, REAC: reactome, WP: WikiPathways, TF: TRANSFAC, MIRNA: miRTarBase, HPA: human protein atlas, HP: human phenotype.

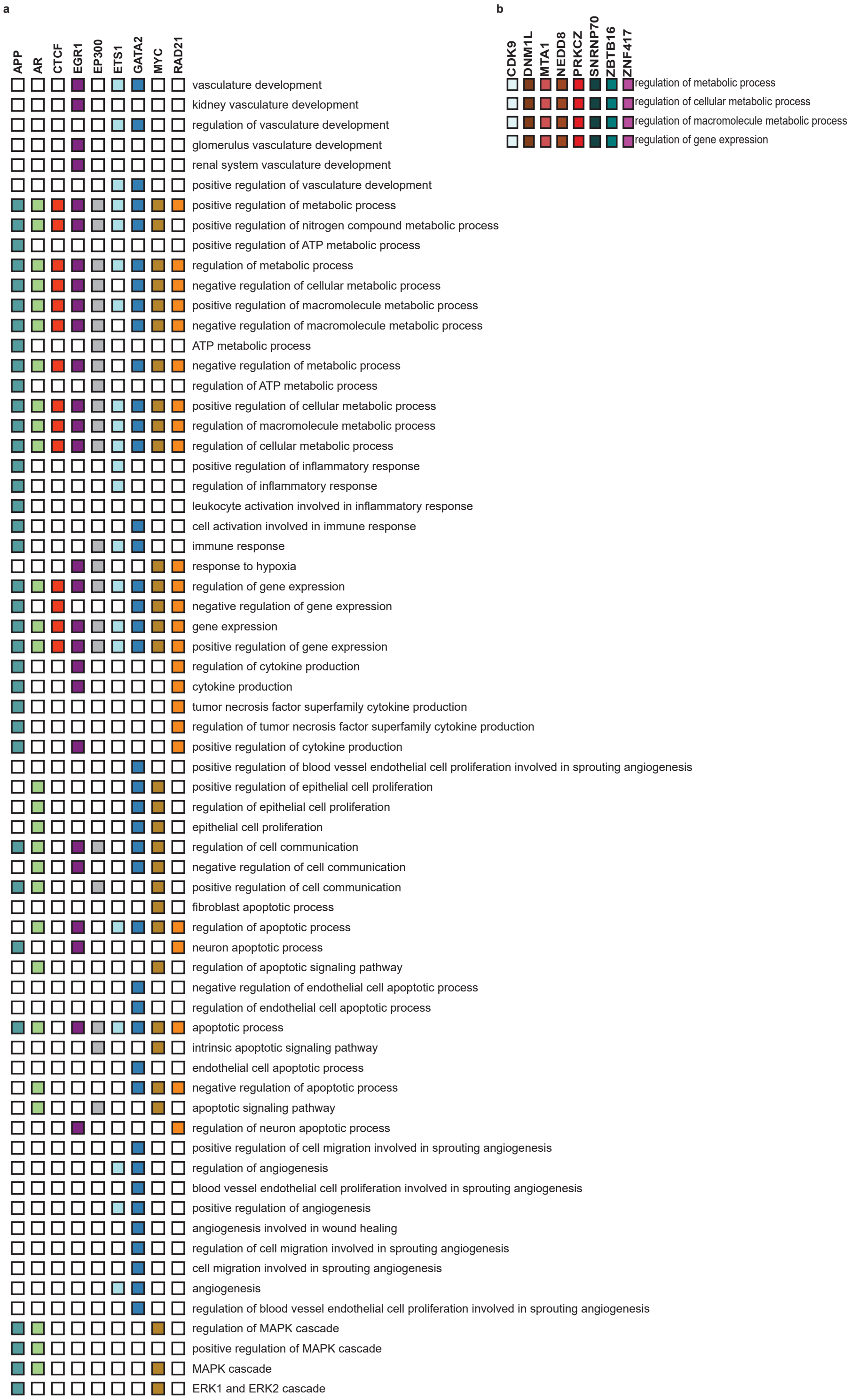


**Supplementary Figure 2. The enriched pathways using the top predicted genes in heart failure**. **a:** is the enriched critical pathways for heart failure (HF) using the predicted genes after integrating the CMR features in the knowledge graph (KG). Only the top nine predicted genes were significantly associated with critical pathways for HF. **b:** is the enriched critical pathways for HF using the predicted genes after excluding the CMR features from the (KG). Only the top eight predicted genes were significantly associated with critical pathways for HF.

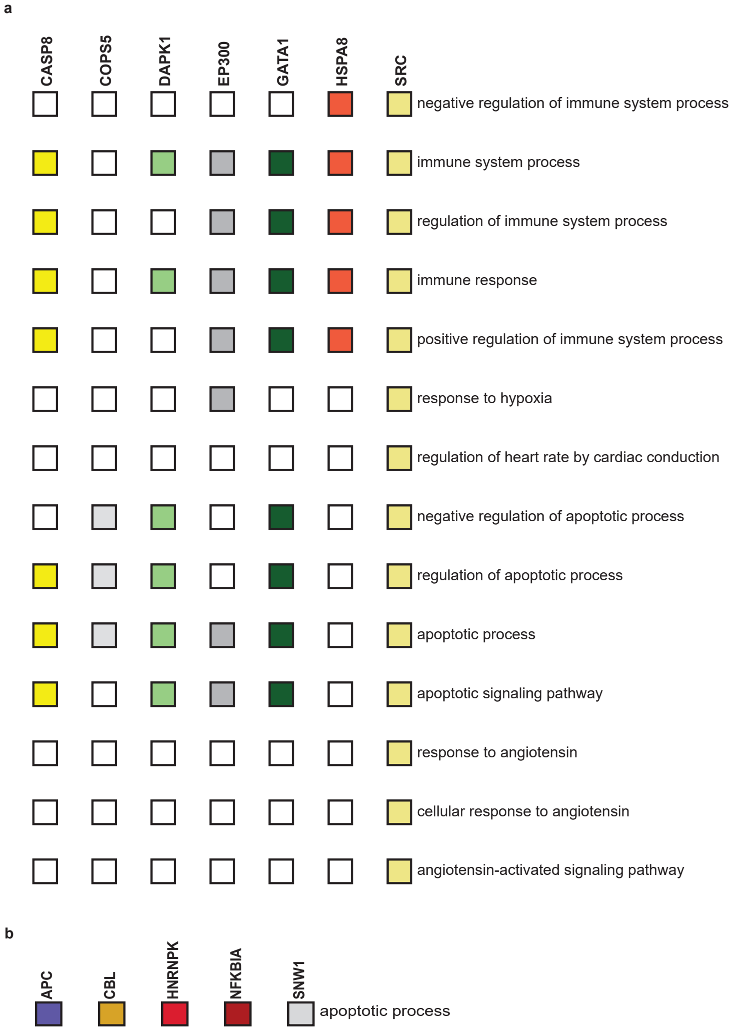


**Supplementary Figure 3. The enriched pathways using the top predicted genes in atrial fibrillation**. **a:** is the enriched critical pathways for atrial fibrillation (AF) using the predicted genes after integrating the CMR features in the knowledge graph (KG). Only the top seven predicted genes were significantly associated with critical pathways for AF. **b:** is the enriched critical pathway for atrial fibrillation using the predicted genes after excluding the CMR features from the (KG). Only the top five predicted genes were significantly associated with critical pathways for AF.

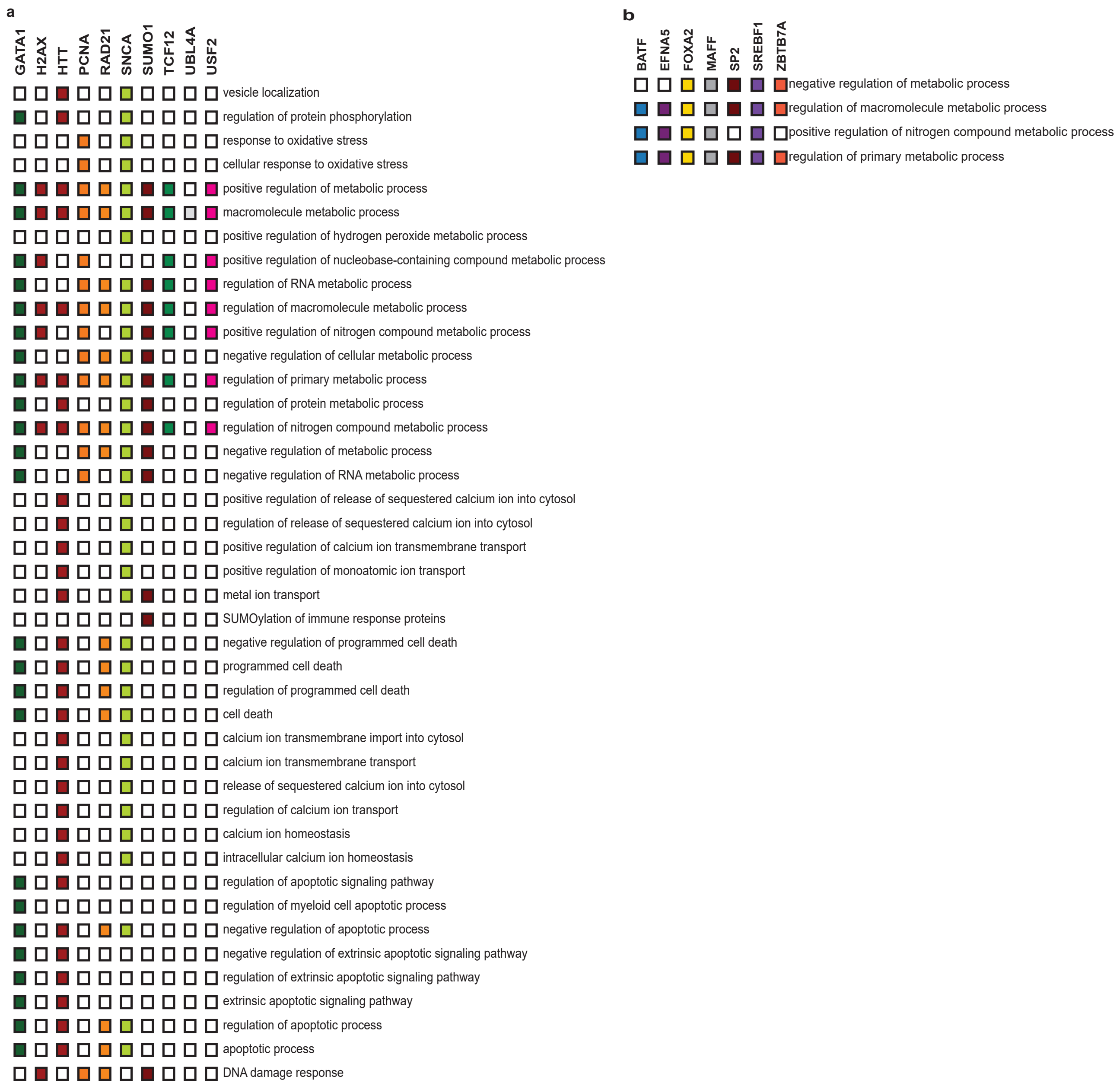


**Supplementary Figure 4. The enriched pathways using the top predicted genes in myocardial infarction**. **a:** is the enriched critical pathways for myocardial infarction (MI) using the predicted genes after integrating the CMR features in the knowledge graph (KG). The top ten predicted genes were significantly associated with critical pathways for MI. **b:** is the enriched critical pathway for myocardial infarction using the predicted genes after excluding the CMR features from the (KG). The top seven predicted genes were significantly associated with critical pathways for MI.

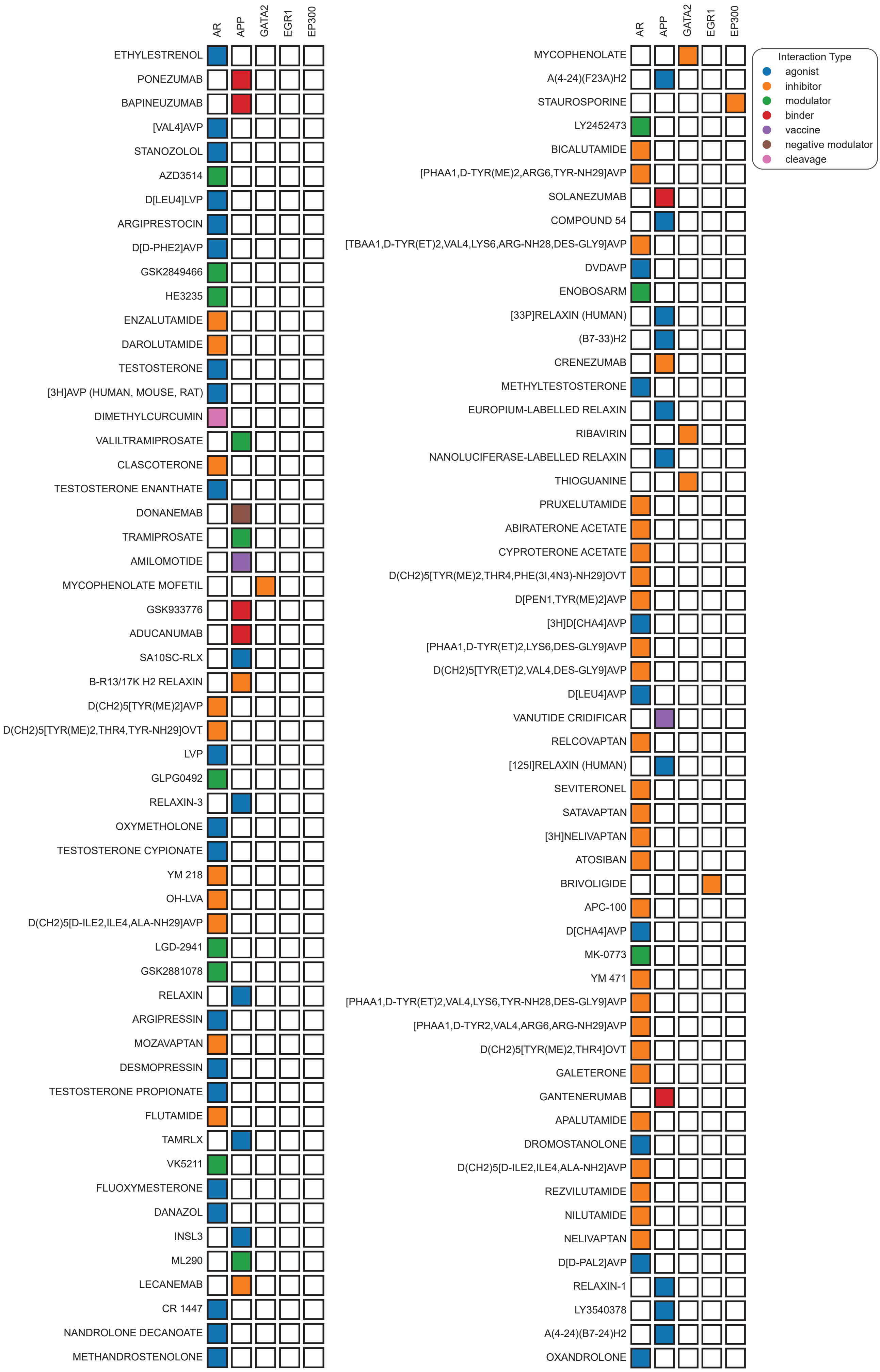


**Supplementary Figure 5. The druggable genes in heart failure with CMR**. Rows names represent medications and columns names represent genes.

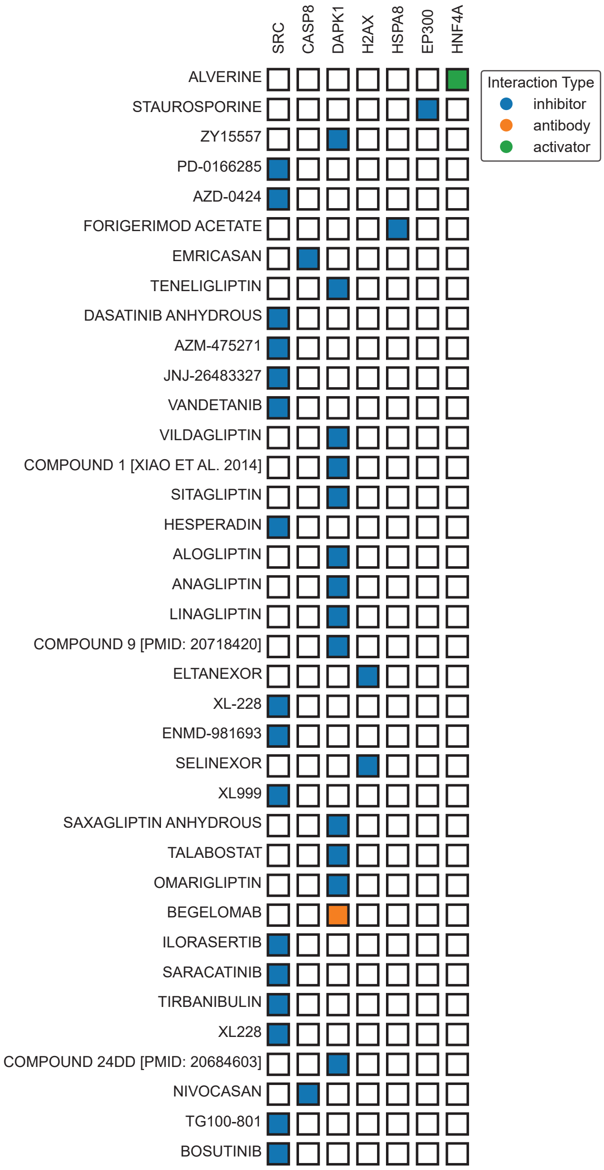


**Supplementary Figure 6. The druggable genes in atrial fibrillation with CMR**. Rows names represent medications and columns names represent genes.

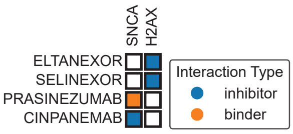


**Supplementary Figure 7. The druggable genes in myocardial infarction with CMR**. Rows names represent medications and columns names represent genes.

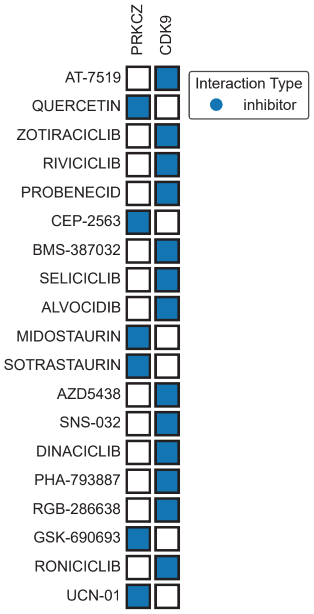


**Supplementary Figure 8. The druggable genes in heart failure without CMR**. Rows names represent medications and columns names represent genes.

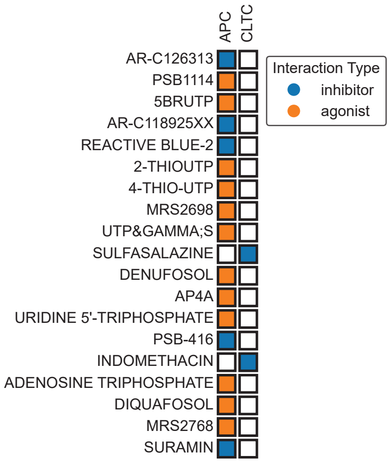


**Supplementary Figure 9. The druggable genes in atrial fibrillation without CMR**. Rows names represent medications and columns names represent genes.

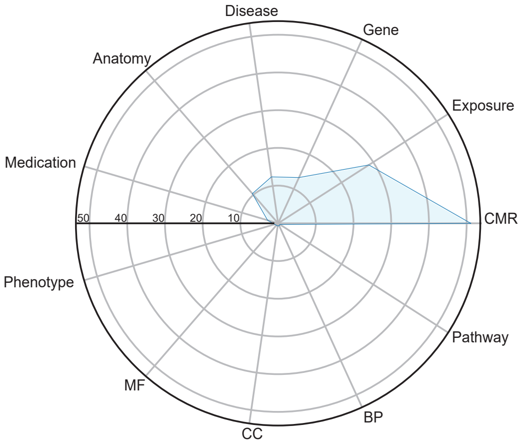


**Supplementary Figure 10. The importance level of the entities in the knowledge graph (KG)**. The radar plot shows the PageRank scores for the entities in the KG. CMR: cardiac magnetic resonance, MF: molecular function, BP: biological process, CC: cellular component.

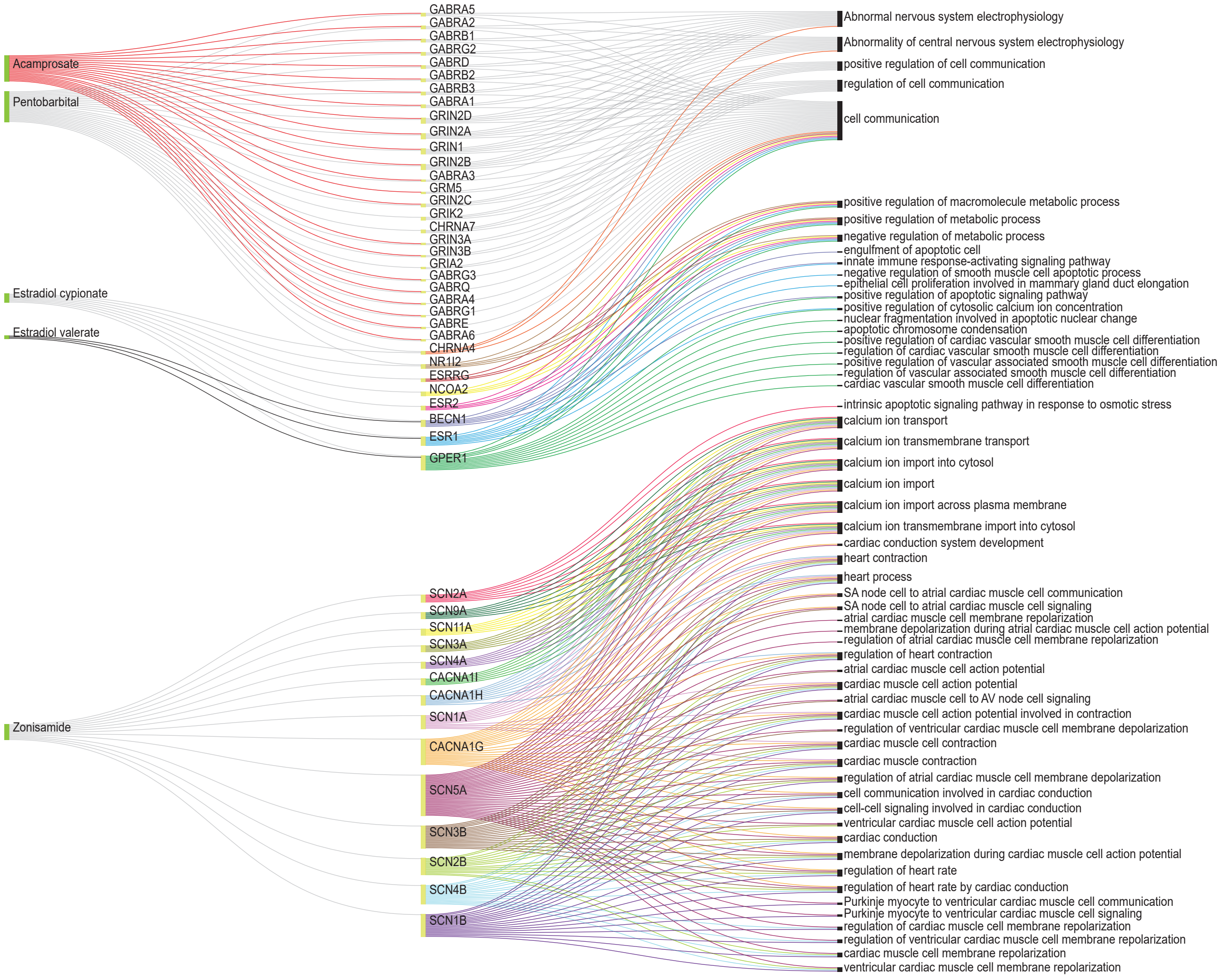


**Supplementary Figure 11. Enrichment analysis results of target genes of predicted medications for atrial fibrillation**. This figure displays the results for pharmaceutical compounds (n=5) only, excluding non-pharmaceutical substances (n=5).

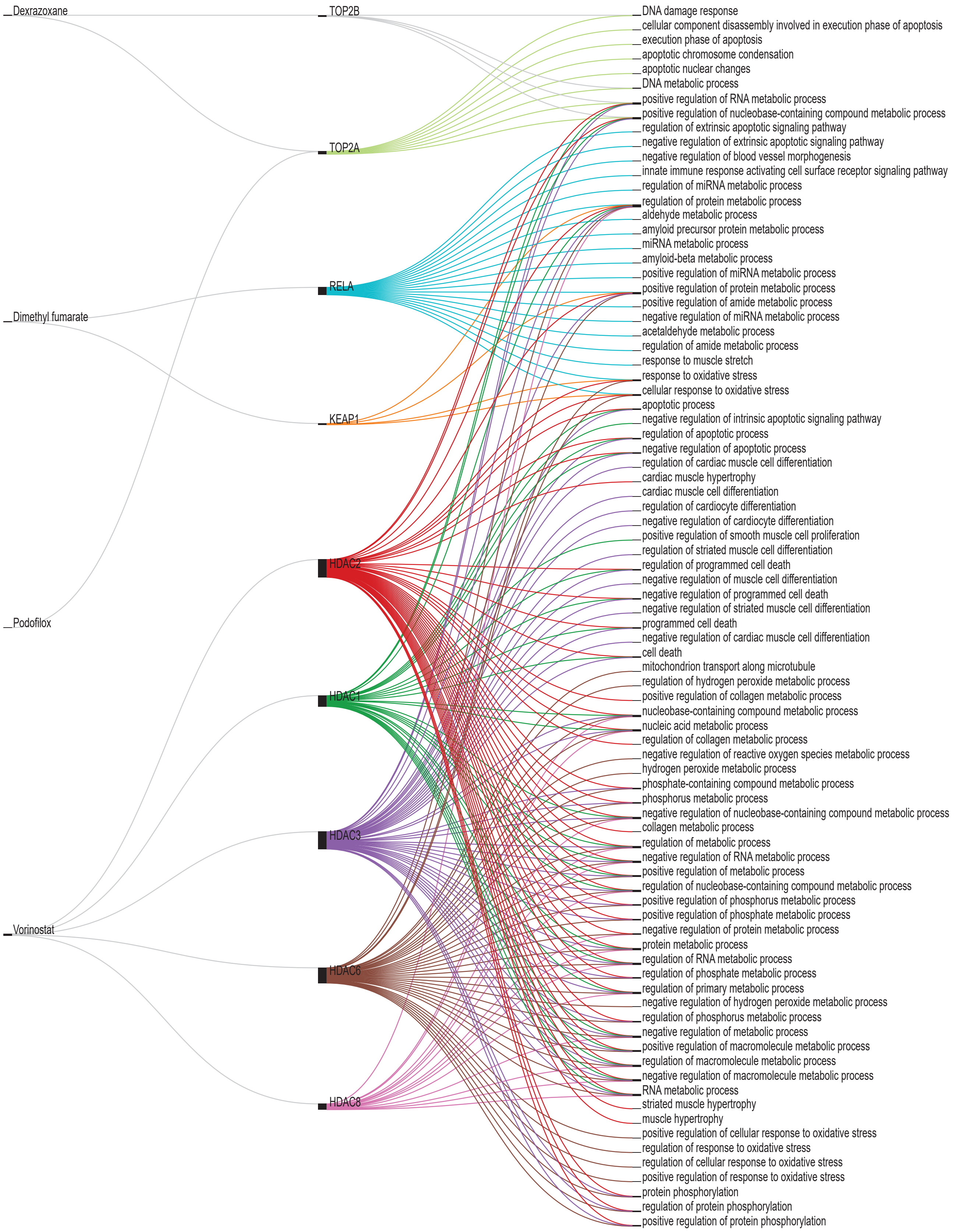


**Supplementary Figure 12. Enrichment analysis results of target genes of predicted medications for myocardial infarction**. This figure displays the results for the first four pharmaceutical compounds (n=4) only.

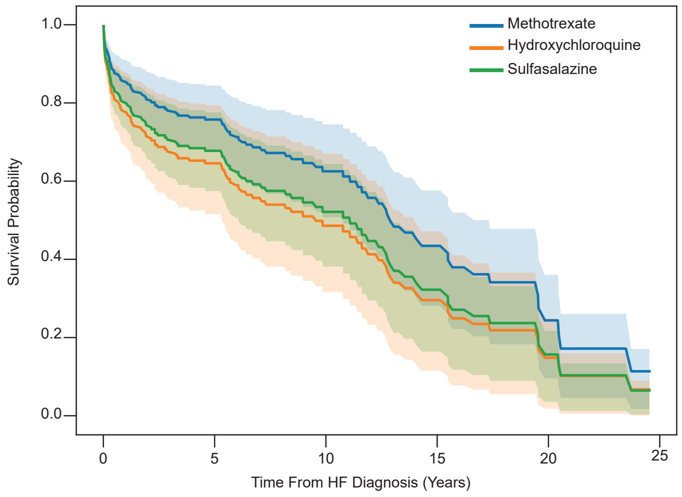


**Supplementary Figure 13. Cox proportional hazards model–based survival curves for Methotrexate (n=147), Hydroxychloroquine (n=47), and Sulfasalazine (n=40)**. The baseline cohort consisted of patients diagnosed with both heart failure (HF) and rheumatoid arthritis (RA). Survival functions were adjusted for age, sex, BMI, and ethnicity. Shaded areas represent 95% confidence intervals. A statistically significant difference in survival was observed between the Methotrexate and Hydroxychloroquine groups (p <0.02)

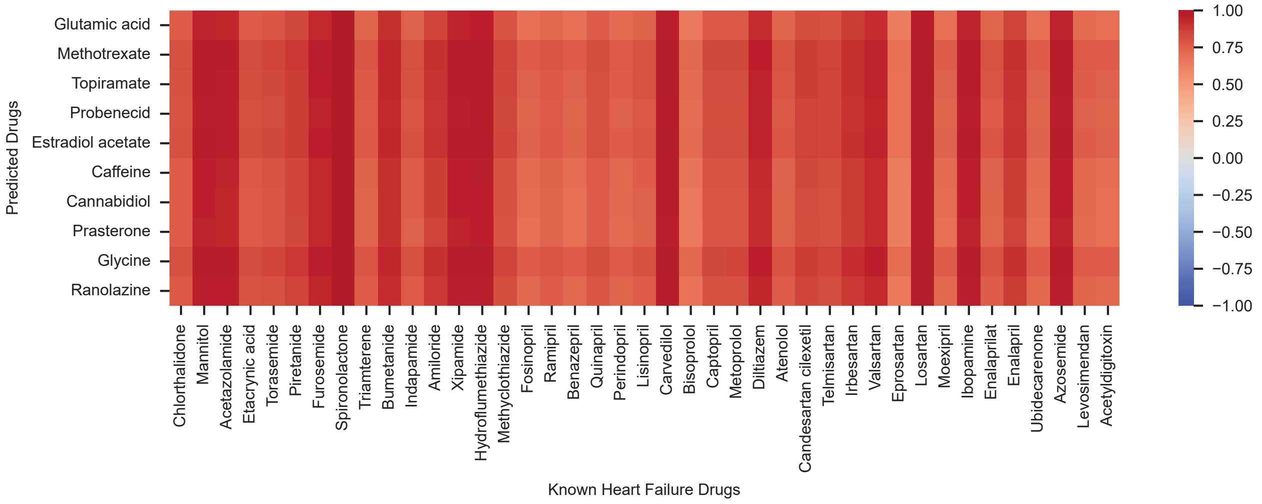


**Supplementary Figure 14. Graph-based validation of the model-predicted medications for heart failure (HF) using embedding similarity analysis**.The figure shows the cosine similarity scores between the predicted medications (based on the knowledge graph) and medications already indicated for HF in clinical practice. Higher cosine similarity indicates greater proximity in the embedding space, suggesting stronger functional or therapeutic relevance. Medications with higher similarity scores are more closely aligned with known HF treatments in the embedding space.

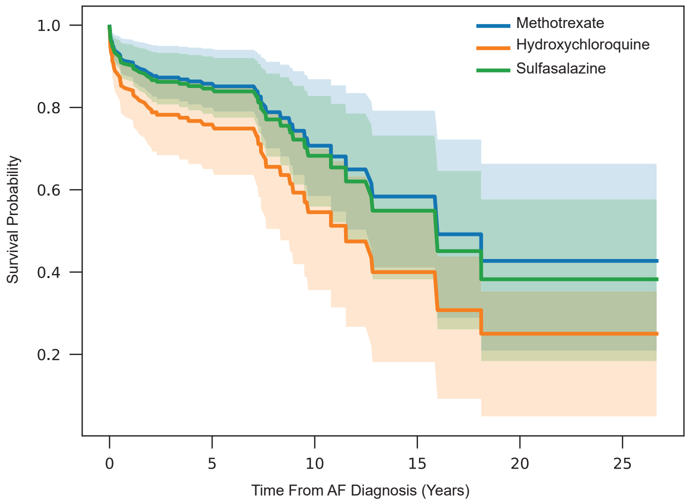


**Supplementary Figure 15. Cox proportional hazards model–based survival curves for Methotrexate (n=170), Hydroxychloroquine (n=39), and Sulfasalazine (n=46)**. The baseline cohort consisted of patients diagnosed with both atrial fibrillation (AF). Survival functions were adjusted for age, sex, BMI, and ethnicity. Shaded areas represent 95% confidence intervals..

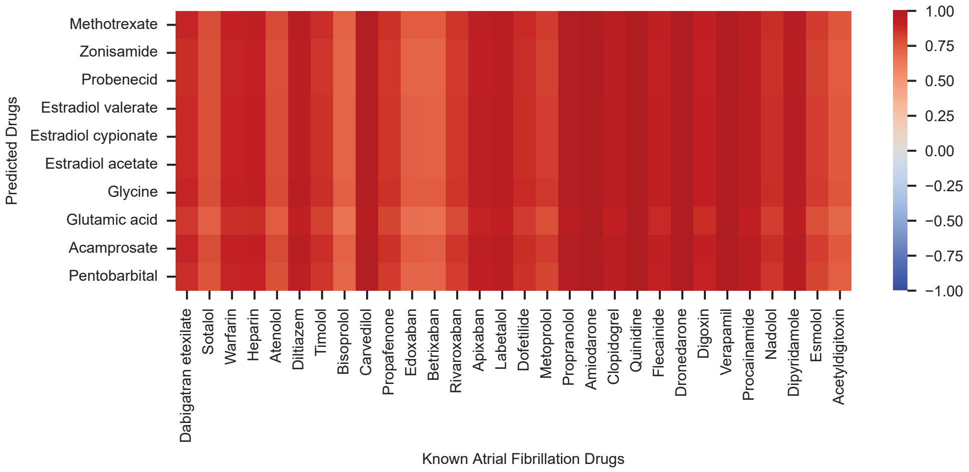


**Supplementary Figure 16. Graph-based validation of the model-predicted medications for atrial fibrillation (AF) using embedding similarity analysis**. The figure shows the cosine similarity scores between the predicted medications (based on the knowledge graph) and medications already indicated for AF in clinical practice. Higher cosine similarity indicates greater proximity in the embedding space, suggesting stronger functional or therapeutic relevance. Medications with higher similarity scores are more closely aligned with known AF treatments in the embedding space.

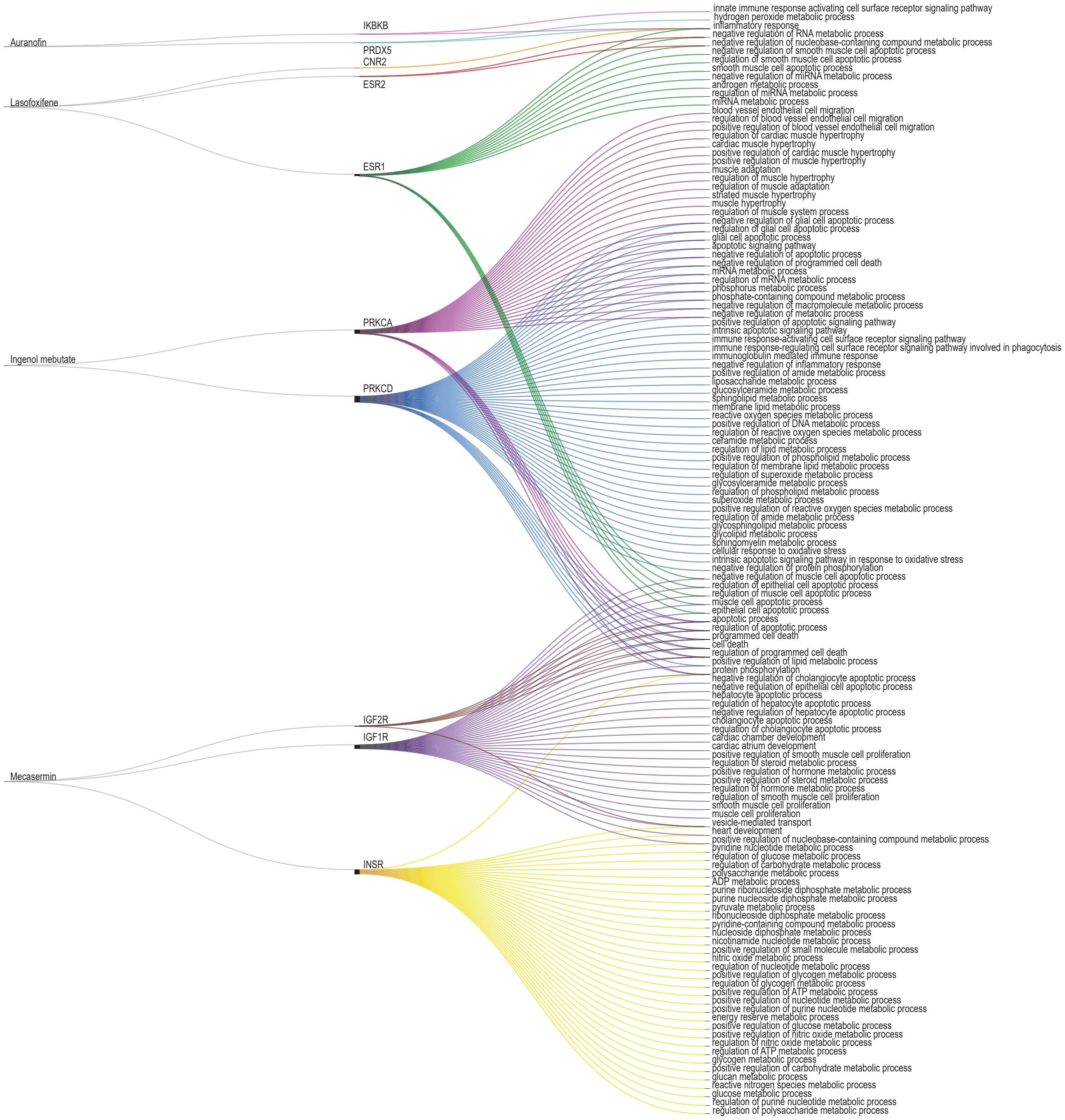


**Supplementary Figure 17. Enrichment analysis results of target genes of predicted medications for myocardial infarction**. This figure displays the results for the next four pharmaceutical compounds (n=4) only.

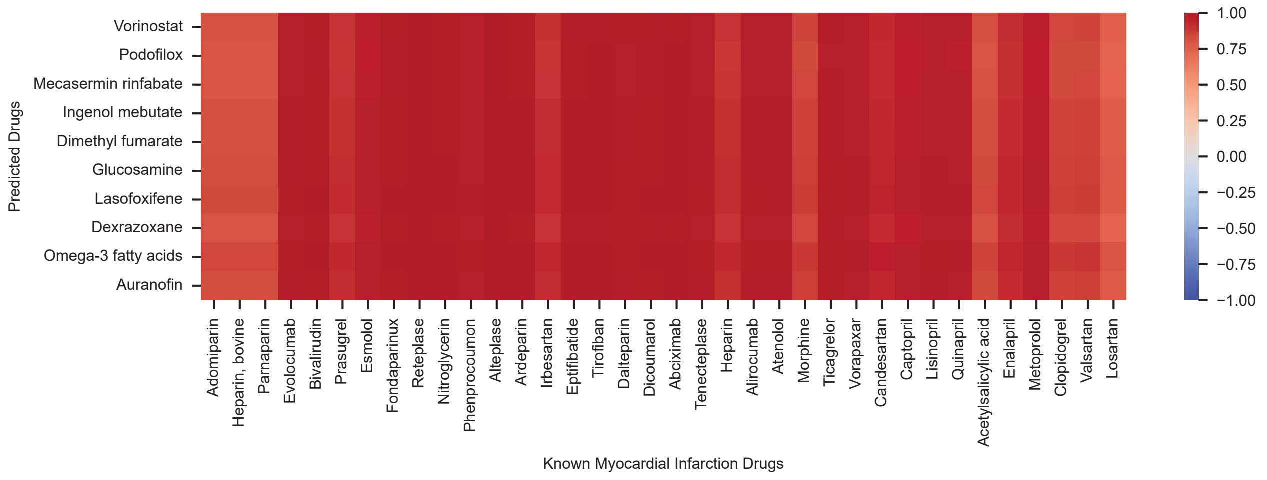


**Supplementary Figure 18. Graph-based validation of the model-predicted medications for myocardial infarction (MI) using embedding similarity analysis**. The figure shows the cosine similarity scores between the predicted medications (based on the knowledge graph) and medications already indicated for MI in clinical practice. Higher cosine similarity indicates greater proximity in the embedding space, suggesting stronger functional or therapeutic relevance. Medications with higher similarity scores are more closely aligned with known MI treatments in the embedding space.

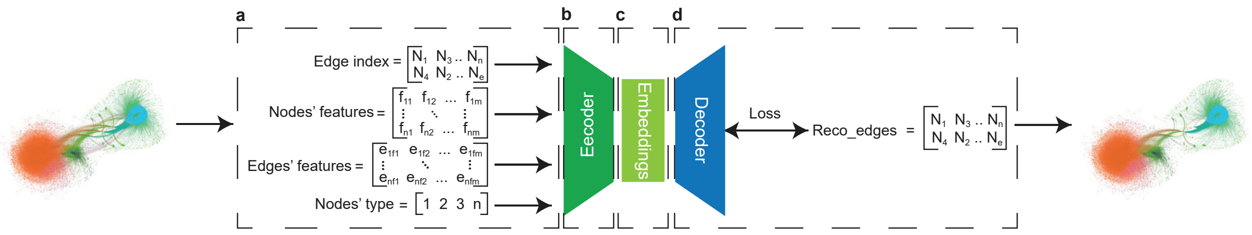


**Supplementary Figure 19. The structure of the Embedding algorithm**. The directed variational graph auto-encoder (DVGAE) operates on four key inputs: edge directions, node properties, relationship attributes, and node types. The encoder processes these inputs to generate embeddings in a latent space, while the decoder reconstructs the original graph to assess the embedding algorithm's performance and the quality of the generated embeddings. These latent-space embeddings are subsequently utilized for the downstream task of gene-disease association prediction. **a**: the key input (matrices and vectors). **b**: the encoder, which takes the graph as four key inputs. **c**: the latent space which has the generated embeddings from the encoder. **d**: the decoder with use the the embeddings to reconstruct the edges and evaluate the quality of the generated embeddings.
